# Assessment of quality of life of breast cancer patients attending a tertiary hospital in Bangladesh

**DOI:** 10.1101/2022.12.02.22283032

**Authors:** Kazi Faria Islam, Abdul Awal, Farzana Tamanna Ummey Shaon, Md. Badrul Hossain, Aliayah Samson, James Senjeh Momo, Mehedi Hasan, Abu. A. M. Hanif, Ilias Mahmud, Malay Kanti Mridha

## Abstract

**Objective:** Breast cancer is the most commonly diagnosed malignancy and a leading cause of death among women. This cross-sectional study assessed the quality of life (QOL) of breast cancer patients attending a tertiary hospital in Dhaka, Bangladesh.

**Method:** Data were obtained from 359 female breast cancer patients at a tertiary hospital between November 23 and December 9, 2019, using a digital platform (Kobo Toolbox). A Bangla translation of the QOL questionnaire (EORTC-QLQ-C30) was used to assess QOL. The survey comprised three scales: global health, functional, and symptoms scale. For the functional and global health scales, we adopted a cut-off level of ≥66% score for good QOL and ≤33% for poor QOL, which was reversed for the symptoms scale. Using STATA 13.0, we performed descriptive and logistic regression analyses.

**Result:** Among the 359 patients, 86.35% were housewives, and 50.42% of them came from the Dhaka division. In contrast to the 38.44% and 42.90 % of respondents who scored highly in the social and cognitive categories of the functional scale, respectively, only 8.92% of patients had a high global health status score. Financial difficulties (28.97%) and fatigue (17.82%) were reported as the most distressing factors followed by loss of appetite (14.76%) and insomnia (13.65%). The logistic regression models revealed that women ≥40 years had higher odds of exhibiting the poor quality of health concerning physical function (AOR 3.59, p=0.005), role function (AOR 3.89, p=0.002), and emotional function (AOR, 2.87, p=0.009) as compared to women<40 years.

**Conclusion:** Our study emphasizes the necessity of integrating psychosocial components for both patients and informal caregivers in the cancer treatment service. Additionally, it is critical to design and implement multi-sectoral policies and integrative techniques to alleviate the most distressing issues for breast cancer patients, as demonstrated in our study: financial hardships, exhaustion, and pain.

## 1. Introduction

Cancer is one of the leading causes of non-communicable diseases (NCDs), which account for the majority of fatalities worldwide and viewed as a significant barrier to the rise in global life expectancy (1). According to estimates, 1.7 million new cases of breast cancer were detected in women globally in 2012, making it the most deadly and prevalent cancer among them (2). Around 60% of new cases of cancer are found in low-income countries (LMIC) in Asia and Africa, which account for 70% of all cancer fatalities worldwide (3). The National Cancer Registry, which is incomplete, is the primary source of information about cancer morbidity in the majority of LMIC, hence the actual incidence of cancer cases is still understated (4). In the coming decades, regardless of available resources, cancer, which is now the top cause of death in many high-income countries, is anticipated to overtake other diseases as a major cause of morbidity and mortality worldwide (5). The combination of negative changes in dietary, reproductive, metabolic, hormonal, and other behavioural aspects is probably what is causing this global increase (1).

Bangladesh is seeing a rapid epidemiological transition from communicable diseases to non-communicable diseases (NCDs), which now account for 61% of the nation’s annual mortality (6). This is a trend that is occurring in many developing countries(7). Among such NCDs, it is projected that 30,000 women each year in Bangladesh pass away from breast cancer, and the mortality rate is rising at a startling rate (2, 8). Recent data from GLOBOCAN, 2018 estimated that 6,846 people died from breast cancer in Bangladesh, which accounted for 19% (12,764) of all new cancer cases among women of reproductive age (5, 7). Women with breast cancer are living longer in high-income nations, while in low and middle-income countries (LMIC), the survival rate is only 10–25% (9, 10).

All cancer patients, regardless of age, may have similar worries, perceptions, fears, and anxieties. However, throughout the past three decades, for women with breast cancer, marriage, having children, getting a career, and finishing education have had an impact on their lives (11–14). A study from Nepal identified critical QOL issues of women with breast cancer referred to as the six Ds—fear of death; dependence on family, spouse, and medical professionals; disfigurement and changes in appearance and self-image with the possibility of endangered sexual and reproductive function, possible early and abrupt menopause; disability making it difficult to complete age-appropriate tasks at work, school; disrupted interpersonal relationships, and discomfort in stages of illness (15).

Depression, anxiety, exhaustion, and psychological trauma not only impair a patient’s quality of life (QOL), but also make it more difficult for them to adhere to anticancer treatments, which has a negative impact on their prognosis, length of stay in the hospital, and even their mortality (14, 16–18). According to a cross-sectional study done in Nepal, depression symptoms were present in 28% of cancer patients, whereas anxiety symptoms were present in 40% of the patients (15). D Women with breast cancer who felt unsupported found it harder to communicate their demands to loved ones and medical professionals (13, 19). These elements may make it difficult for individuals to get the social assistance they need, which increases their risk of psychosocial and psychosexual illness and lowers their quality of life overall (12). An additional key issue affecting the patient’s welfare as well as the welfare of future generations of the patient’s family, according to a study from Punjab, India, is the catastrophic health expenditure brought on by out-of-pocket (OOP) payments for cancer diagnosis and treatment (20). Additionally, the socioeconomic burden, worry about a recurrence, and even death; are all of these factors directly or indirectly connected to the quality of life of women with breast cancer (11, 21).

The likelihood of long-term cancer-free survival among women has increased because of early detection of breast cancer, comprehensive adjuvant therapy, hormone therapy, and conservative treatment (22–24). A crucial factor to take into account is the psychological distress level experienced by women during the entire process, from screening to receiving treatment (25). Since the World Health Organization expanded the definition of health to include physical, mental, and social well-being in 1948 (26), health-related QOL issues have become a significant aspect of healthcare practice and research (24, 27). Literature suggest that a person’s health-related quality of life is influenced by his or her survival experiences, values, expectations, and perceptions (HRQL) (28, 29). For the past 20 years, HRQL has been significantly used as a tool to forecast and assess patient outcomes and treatment interventions (30).

To understand the effects of any chronic disease, doctors and policymakers are finally realizing the relevance of measuring health-related quality of life (HRQL) (31–33). In South Asia, studies on the quality of life for young women with breast cancer were carried out in Nepal (34), Sri Lanka (35), and South India (36). But as far as we are aware, no research has been done in Bangladesh (6). A study focusing on the perception of social support, the vulnerability of the cohort, the conundrum, and quality of life among younger women with breast cancer in Bangladesh, where few treatment opportunities are available for affordable comprehensive cancer treatment is unquestionably necessary (10, 37). Considering the dearth of data available in Bangladesh and the significance of QOL in the prognosis of cancer treatment the present study assessed the quality of life of female breast cancer patients undergoing treatment at a tertiary hospital in Dhaka, Bangladesh.

## 3. Methodology

The present cross-sectional study was carried out in the Day-care Centre of the National Institute of Cancer Research & Hospital (NICRH), Mohakhali, Dhaka, Bangladesh where patients with cancer come for chemotherapy from 23rd November to 9th December 2019. The study was conducted at NICRH because it provides a comprehensive approach to treating cancer including prevention, diagnosis, treatment, and rehabilitation for a wide spectrum of patients who present with various cancer stages and from diverse age groups.

Participants in the study were to be female patients between the ages of 18 and above who were receiving chemotherapy at the day-care centre and had been diagnosed with breast cancer at any stage. We employed convenience sampling to recruit participants in the survey and excluded patients who were critically ill and admitted to the indoor department. To assess the quality of life of breast cancer patients, a structured questionnaire was created using the EORTC QLQ-C30, a tool developed by the European Organization for Research and Treatment of Cancer (38). This instrument has previously been utilized in numerous investigations with various populations and cancer types. The tool we utilized has 30 questions, including a global health status/QoL scale, three symptom scales (fatigue, nausea/vomiting, and pain), five functional scales (physical, role, cognitive, emotional, and social), and six single items (dyspnoea, insomnia, appetite loss, constipation, diarrhoea, and financial difficulties). In this version of the questionnaire, 28 items were rated on a response scale of “not at all” (1) to “very much” (4). The time frame was “during the past week”. The response choices ranged from “extremely poor” (1) to “excellent” (7) for questions 29 (overall general health) and 30 (on overall QoL), respectively, and the time frame was “within the last week”. We also gathered the respondents’ sociodemographic data and physical features. High scores on functional scales characterize healthy functioning. Likewise, a high score for global health status denotes a higher quality of life. However, high scores on symptom ratings indicate a higher level of issues. Scores on all scales and individual items range from 0 and 100. According to the EORTC module’s scoring manual, we used the same principle while scoring these scales (38).

We also gathered respondents’ sociodemographic data regarding age, religion, education status, employment status, marital status, place of residence, number of kids, income, and medical characteristics.

We calculated the sample size for this study using the following formula,

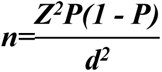

where, n=minimum sample size, Z=confidence interval (95%), P=prevalence of good quality of life among women with breast cancer described by a previous study conducted in Saudi Arabia (54.9%) (39), d= absolute error or precision (5%). The sample size, by using the formula, was estimated to be 380. Keeping the non-participation at 10% the sample size for this study was 418.

We conducted in-person interviews using Kobo Toolbox software through tablet computers. All the researchers were introduced to the day-care centre’s on-duty nurses on the first day of data collection by the on-call doctors. Before conducting interviews researcher introduced herself/himself to each eligible respondent, went over the steps of the study, and provided them with a written consent form. Because our study issue was so delicate, all of the interviewers showed patience and empathy toward the respondents. The team members clarified any questions that the respondents were unclear about. The next eligible respondent was approached to obtain data for any unwilling respondents. Following the completion of each interview, the entire questionnaire was reviewed to ensure consistency, data quality, and completeness.

The ethical review board of the BRAC James P Grant School of Public Health (JPGSPH) reviewed and approved this study with ref no: ID 2019-MPH-SLP 07. Before collecting data, permission was obtained from the National Institute of Cancer Research & Hospital. All respondents provided informed written consent; those who were illiterate provided verbal consent. Throughout the study, the respondents’ privacy and anonymity were upheld. Each questionnaire was given a unique code, and after the study is completed, the data will be stored by JPGSPH policy.

### Statistical analysis

All raw data were entered into the Microsoft Excel database from Kobo Toolbox software which was afterward uploaded to STATA 13.0 for analysis and data cleansing. At the end of our data collection period, data cleaning was done, and any missing values were eliminated from the study. We calculated descriptive statistics for the sociodemographic and medical characteristics.

We calculated the raw score, which is the average of the items that contribute to the scale. To standardize the raw score and convert it to a value between 0 and 100, we first performed a linear transformation. A higher score denotes either a greater (”good”) level of functioning or a higher (”worse”) degree of symptoms. Poor quality of life and good quality of life were the two categories used to determine the quality of life. According to their scores, patients were separated into two groups: those with functional scale and global quality of life scores ≤ 33 were regarded to be in poor condition, while those with scores ≥ 66 were considered to be in good condition. For symptom scales, the score was reversed, indicating that patients scoring 33 or lower were considered to be in good condition and those scoring 66 or lower were regarded to be in poor condition (38, 39). Using STATA 13.0, the data were coded into these categories.

Chi-square test and multivariable logistic regression analyses were done to measure the significance of the association between the dependent and independent variables with significance kept at 95% confidence (p-value <0.05). The dependent variables were global health, physical, emotional, cognitive, and social functioning scores, while age, religion, religion, education status, employment status, marital status, place of residence, number of kids, and menstrual status were the independent variable and considered as the model’s predictor.

## 4. Result

Through face-to-face interviews with our questionnaire, we conducted interviews with 364 female patients diagnosed with breast cancer. Among them, we discarded 5 interviews because of incomplete responses. Socio-demographic characteristics of the 359 breast cancer patients are given below in **Table 1**.

**Table 1.**
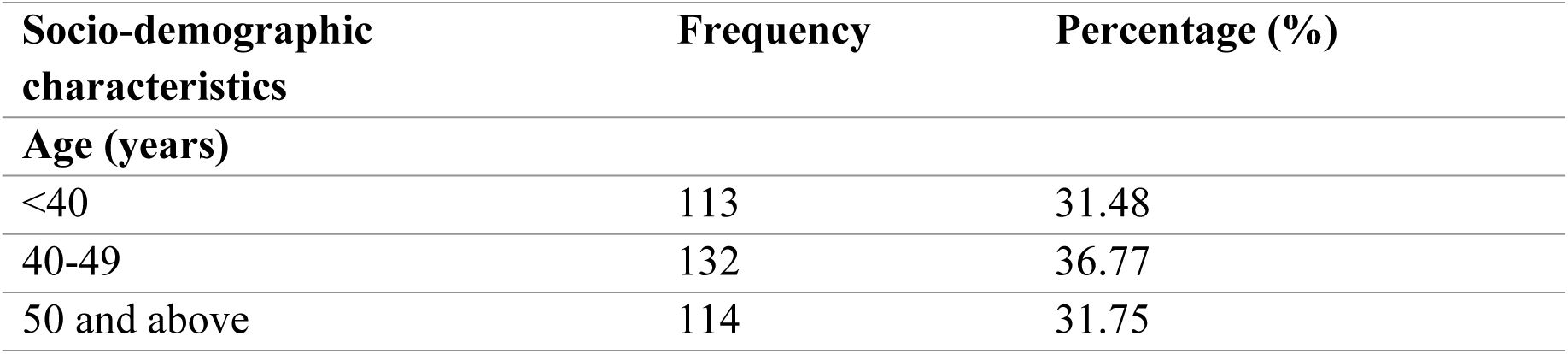
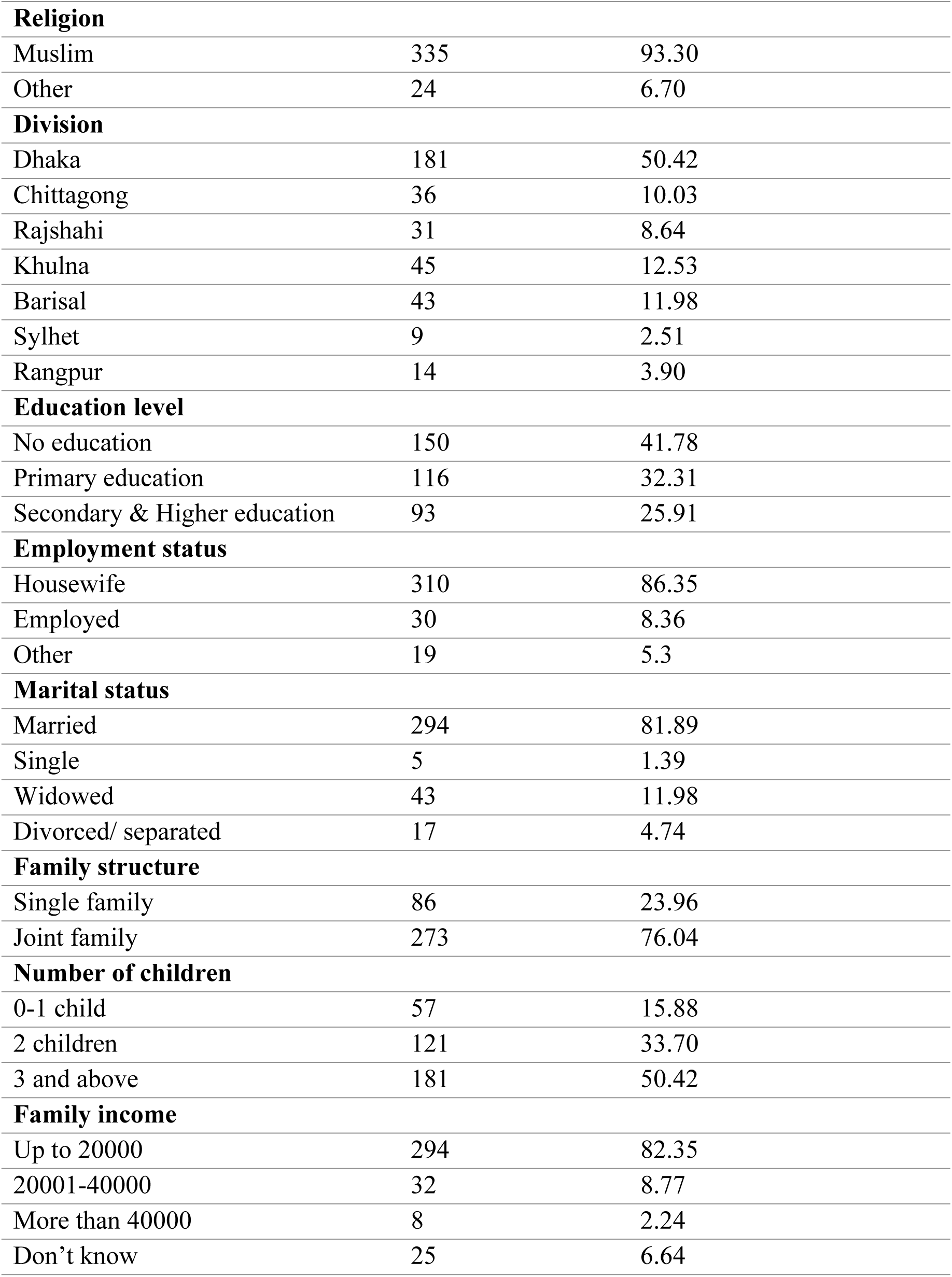
Socio-demographic characteristics of the study subjects (n=359):

| Socio-demographic characteristics | Frequency | Percentage (%) |
| --- | --- | --- |
| Age (years) |  |  |
| <40 | 113 | 31.48 |
| 40-49 | 132 | 36.77 |
| 50 and above | 114 | 31.75 |
| **Religion** |  |  |
| Muslim | 335 | 93.30 |
| Other | 24 | 6.70 |
| **Division** |  |  |
| Dhaka | 181 | 50.42 |
| Chittagong | 36 | 10.03 |
| Rajshahi | 31 | 8.64 |
| Khulna | 45 | 12.53 |
| Barisal | 43 | 11.98 |
| Sylhet | 9 | 2.51 |
| Rangpur | 14 | 3.90 |
| **Education level** |  |  |
| No education | 150 | 41.78 |
| Primary education | 116 | 32.31 |
| Secondary & Higher education | 93 | 25.91 |
| **Employment status** |  |  |
| Housewife | 310 | 86.35 |
| Employed | 30 | 8.36 |
| Other | 19 | 5.3 |
| **Marital status** |  |  |
| Married | 294 | 81.89 |
| Single | 5 | 1.39 |
| Widowed | 43 | 11.98 |
| Divorced/ separated | 17 | 4.74 |
| **Family structure** |  |  |
| Single family | 86 | 23.96 |
| Joint family | 273 | 76.04 |
| **Number of children** |  |  |
| 0-1 child | 57 | 15.88 |
| 2 children | 121 | 33.70 |
| 3 and above | 181 | 50.42 |
| **Family income** |  |  |
| Up to 20000 | 294 | 82.35 |
| 20001-40000 | 32 | 8.77 |
| More than 40000 | 8 | 2.24 |
| Don't know | 25 | 6.64 |

Among the 359 patients interviewed, 36.77% of the respondents were from 40-49 years of age group. Almost all the respondents belonged to the Muslim religion (93.30%). The majority of the respondents (50.42%) came from Dhaka division, while only 2.51 % of respondents were from Sylhet. Around 32% of the respondents had primary education but a major proportion of them (41.78%) reported having no education. Around 82% of our respondents were married. Approximately 76% lived in a joint family and the majority of the respondents (50.42%) had 3 or more children. A large number of respondents (86.35%) were housewives. Among 359 females, 6.64% were not able to report the amount of family income, but 82.35% of the respondents mentioned having an income of up to 20,000 BDT (253.63$) monthly.

### Medical History of the respondents

Respondents were asked to remember the duration since diagnosis of breast cancer. The majority of the respondents (68%) **(Fig 1)** told that their breast cancer was detected in the past 1 year and around 74% **(Fig 2)** of the respondent started their treatment within the 1^st^ year of diagnosis.

**Fig 1.**
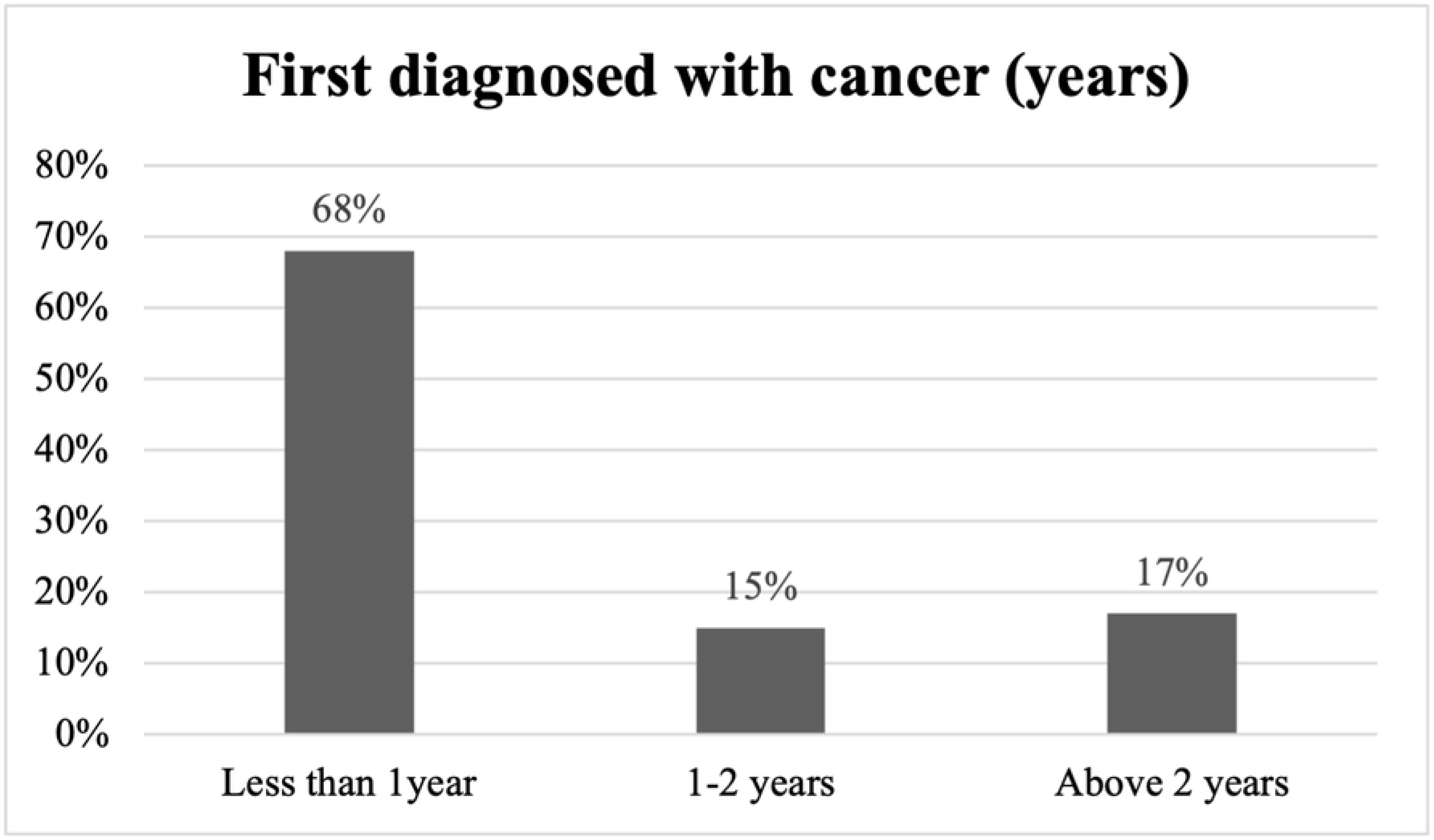
First diagnosed with cancer (years)

**Fig 2.**
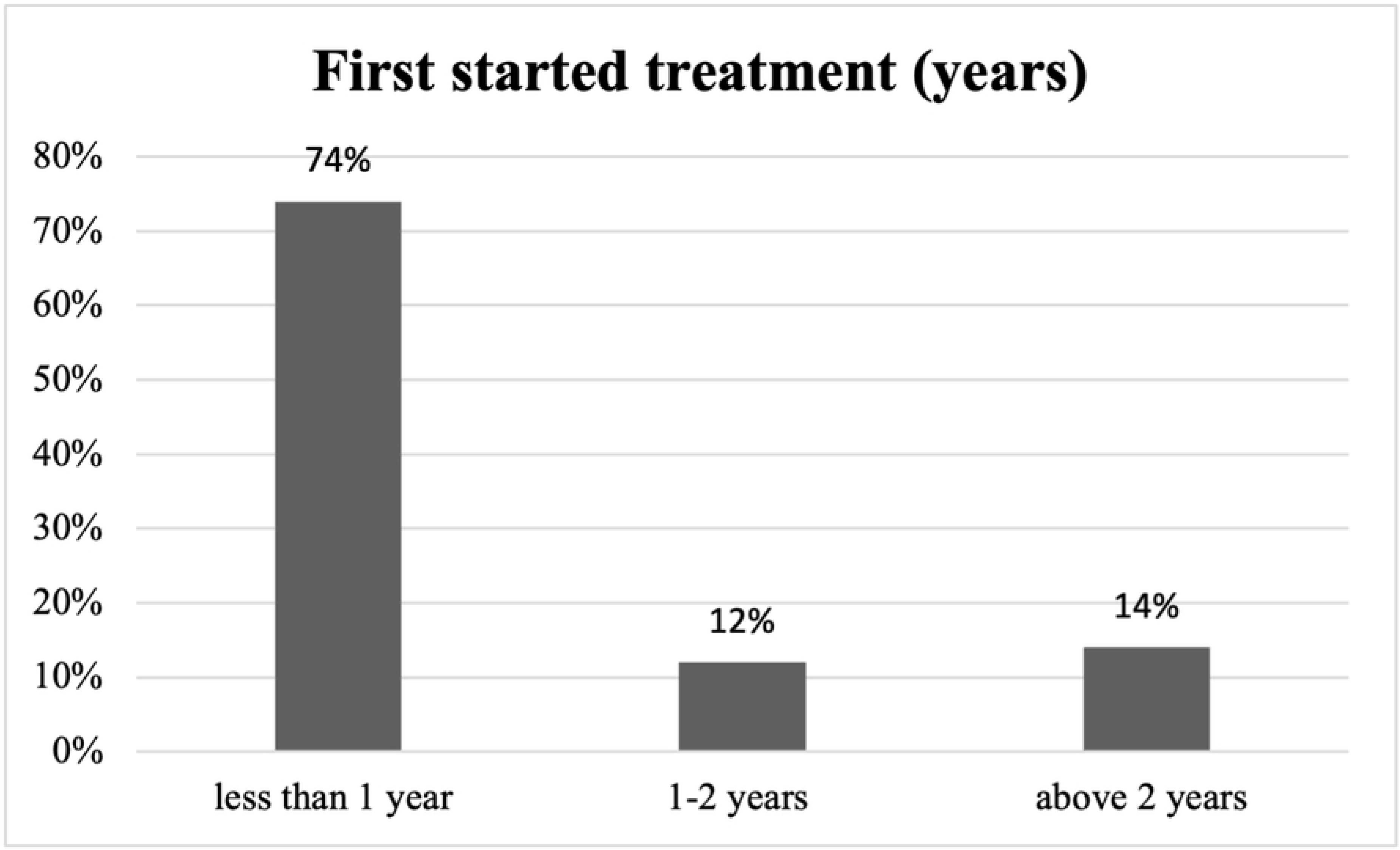
First started treatment (years)

The following table (Table 2) shows that when asked the patients, if they know about the staging of cancer, the majority of the patients (72%) said they did not know their stage of cancer. Only 13% of the respondents could answer that they were in the 2^nd^ stage of cancer. The question of co-morbidity had multiple answers. Only 4% of the respondents said they had other forms of cancer as well, but diabetes (21%) and hypertension (23%) were reported by most of the respondents.

**Table 2.**
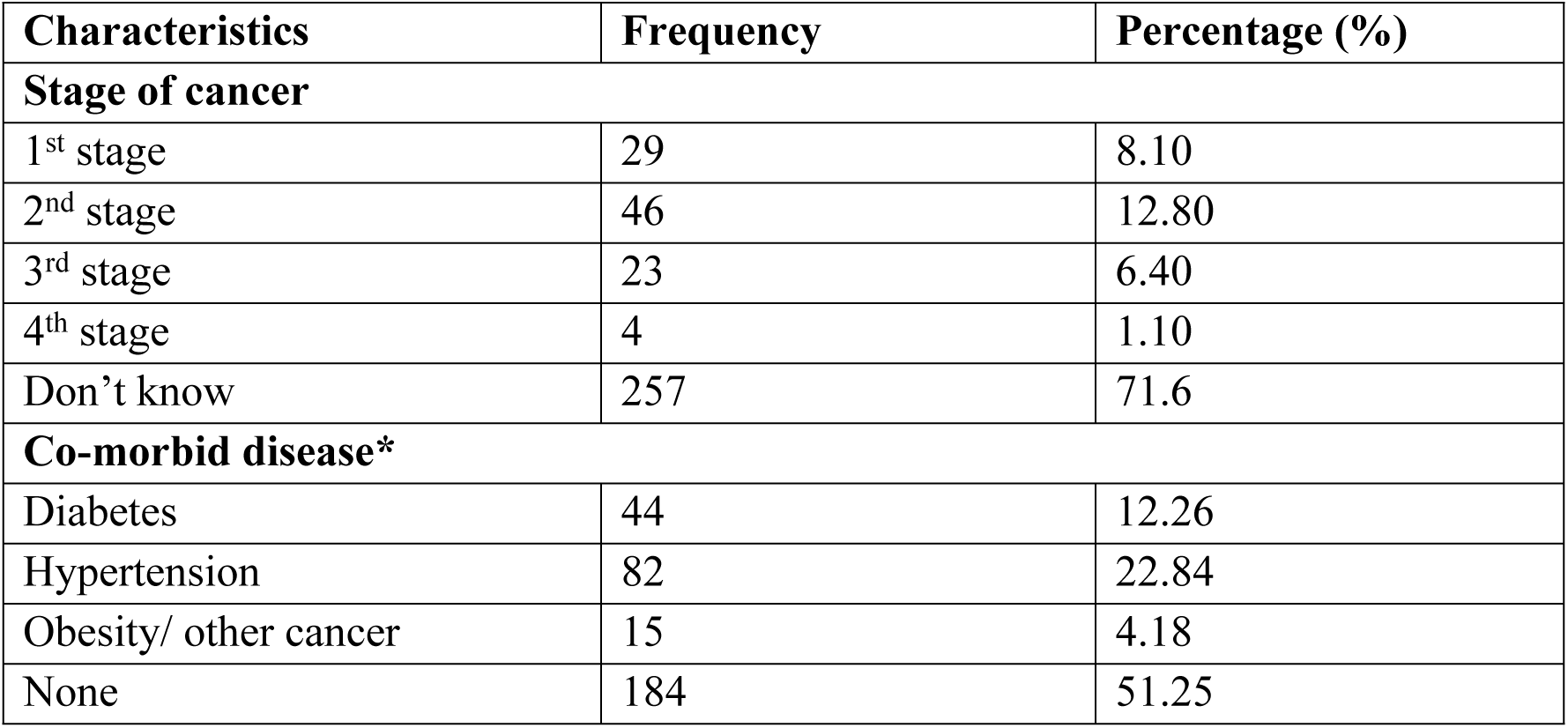
knowledge on stage of cancer and co-morbid disease of female patients.

| Characteristics | Frequency | Percentage (%) |
| --- | --- | --- |
| **Stage of cancer** |  |  |
| 1<sup>st</sup> stage | 29 | 8.10 |
| 2<sup>nd</sup> stage | 46 | 12.80 |
| 3<sup>rd</sup> stage | 23 | 6.40 |
| 4<sup>th</sup> stage | 4 | 1.10 |
| Don't know | 257 | 71.6 |
| **Co-morbid disease\*** |  |  |
| Diabetes | 44 | 12.26 |
| Hypertension | 82 | 22.84 |
| Obesity/ other cancer | 15 | 4.18 |
| None | 184 | 51.25 |

### Type of treatment

When the respondents were asked about the type of treatments they underwent for breast cancer, two-thirds of them (70%) said they underwent both surgery and chemotherapy. Following chart shows only 12% of the respondents took radiotherapy along with surgery and chemotherapy. The following Fig 3 shows it clearly.

**Fig 3.**
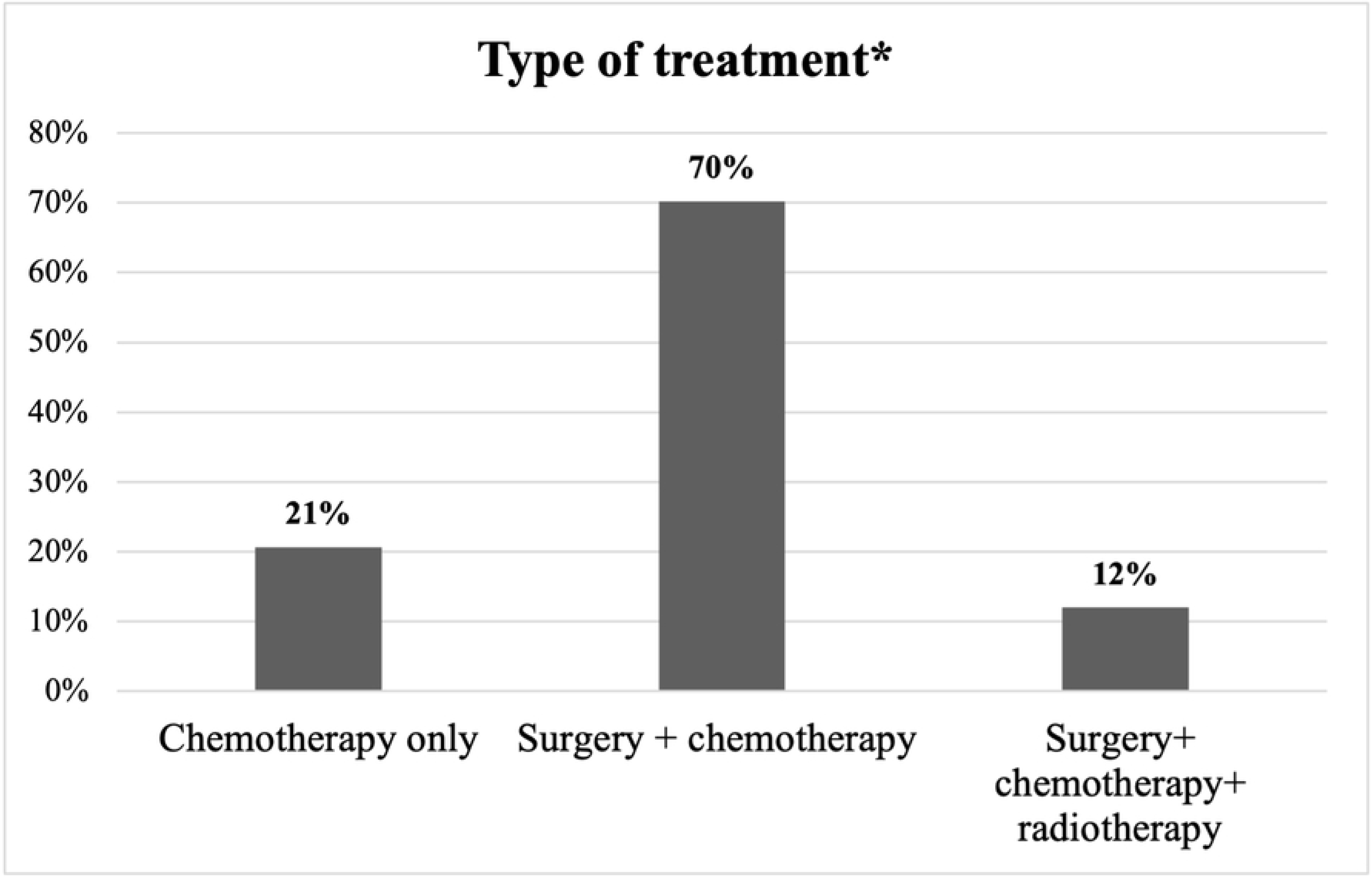
Type of treatment*. *Multiple responses

### Breast Cancer treatment and physical well-being

Literature show treatment for cancer e.g. chemotherapy and radiotherapy affects the menstrual status of women. Among our 359 respondents, around 73% reported that after the treatment their menstruation stopped. Fig **4** summarizes the responses.

**Fig 4.**
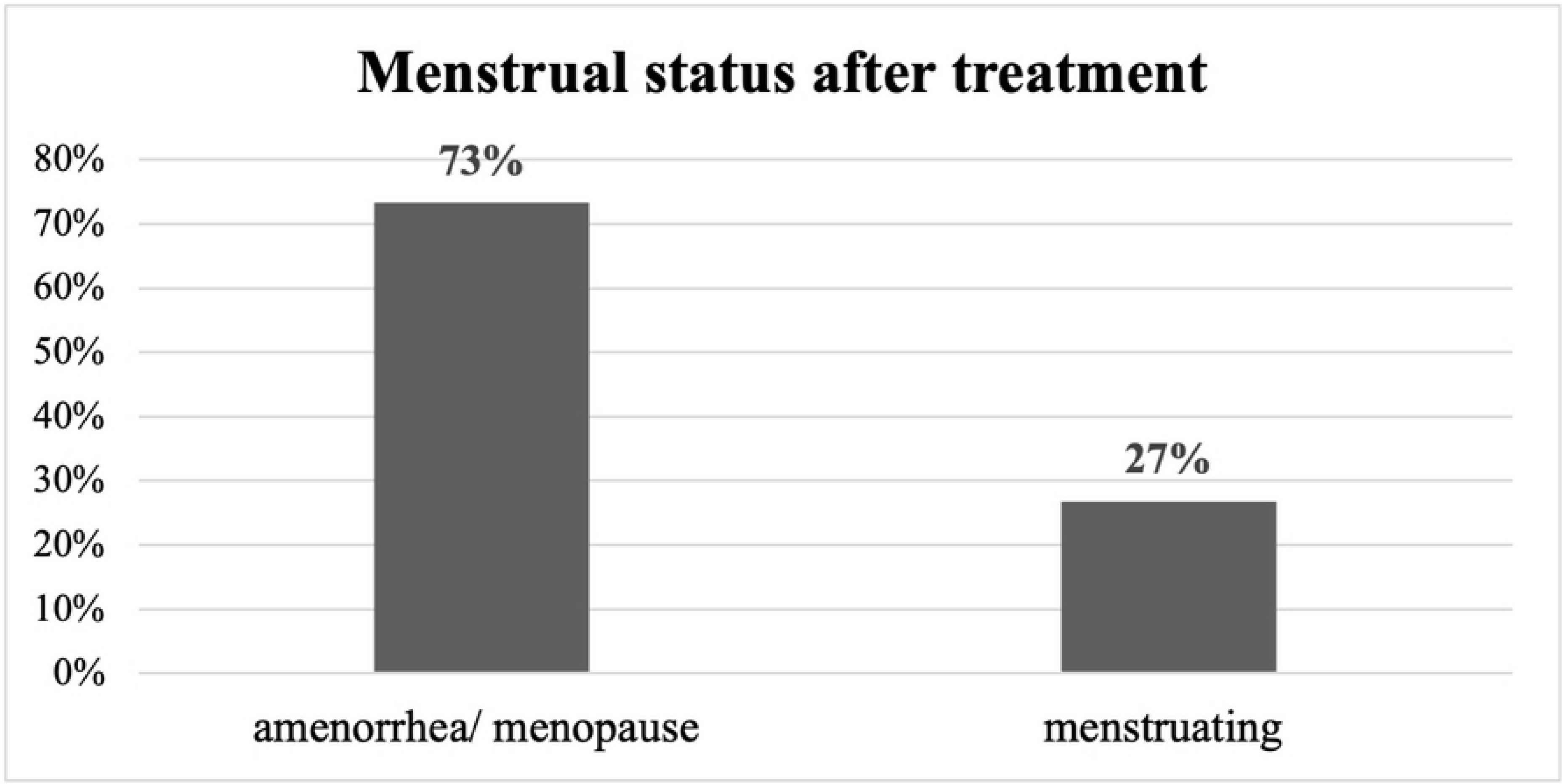
Menstrual status after treatment.

### Pain in stages of cancer

Respondents were questioned about their pain levels in their arms, the condition of their breasts, and their ability to elevate or move their arms sideways at various phases of the disease. The options for each question were “not at all,” “a little,” “quite a bit,” and “very much” and were based on the participants’ last week of memory. The majority of respondents (80%) chose “a little” in response to the questions about discomfort in the arm or shoulder and pain in the affected breast, whereas 39% chose “not at all” in response to the question about difficulty raising the arms or shoulders. The data are shown in **Fig 5.**

**Fig 5:**
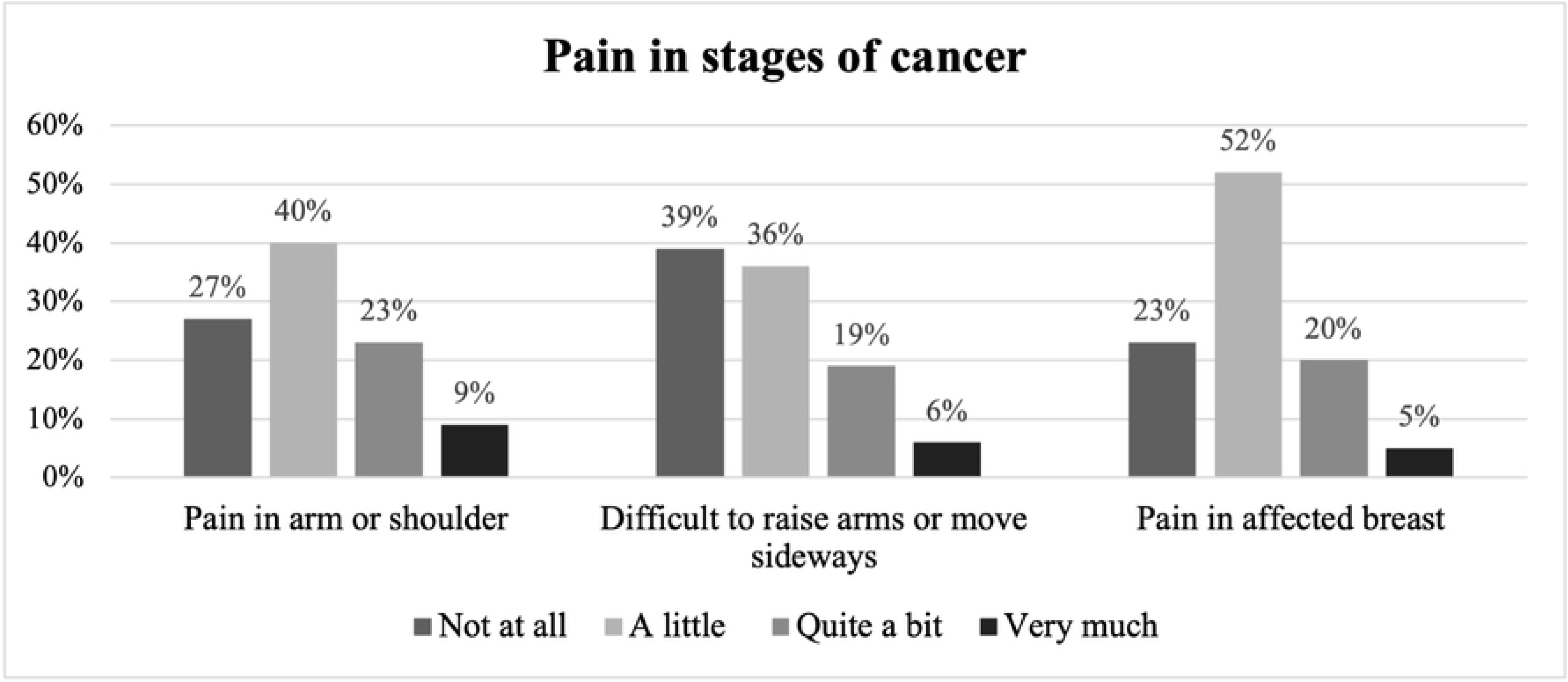
Pain in stages of cancer (%)

### Breast cancer and psychosocial well-being

When asked if they have lost hair as a result of the treatment, the majority of respondents (57%) said “very much.” Nearly 36% of the patients reported feeling extremely melancholy as a result of their hair loss, and 28% stated they felt less attractive, but nearly 37% claimed they were “not at all” unhappy with their bodies. **Fig 6** shows specifics of the reactions related to physical appearance and self-image.

**Fig 6.**
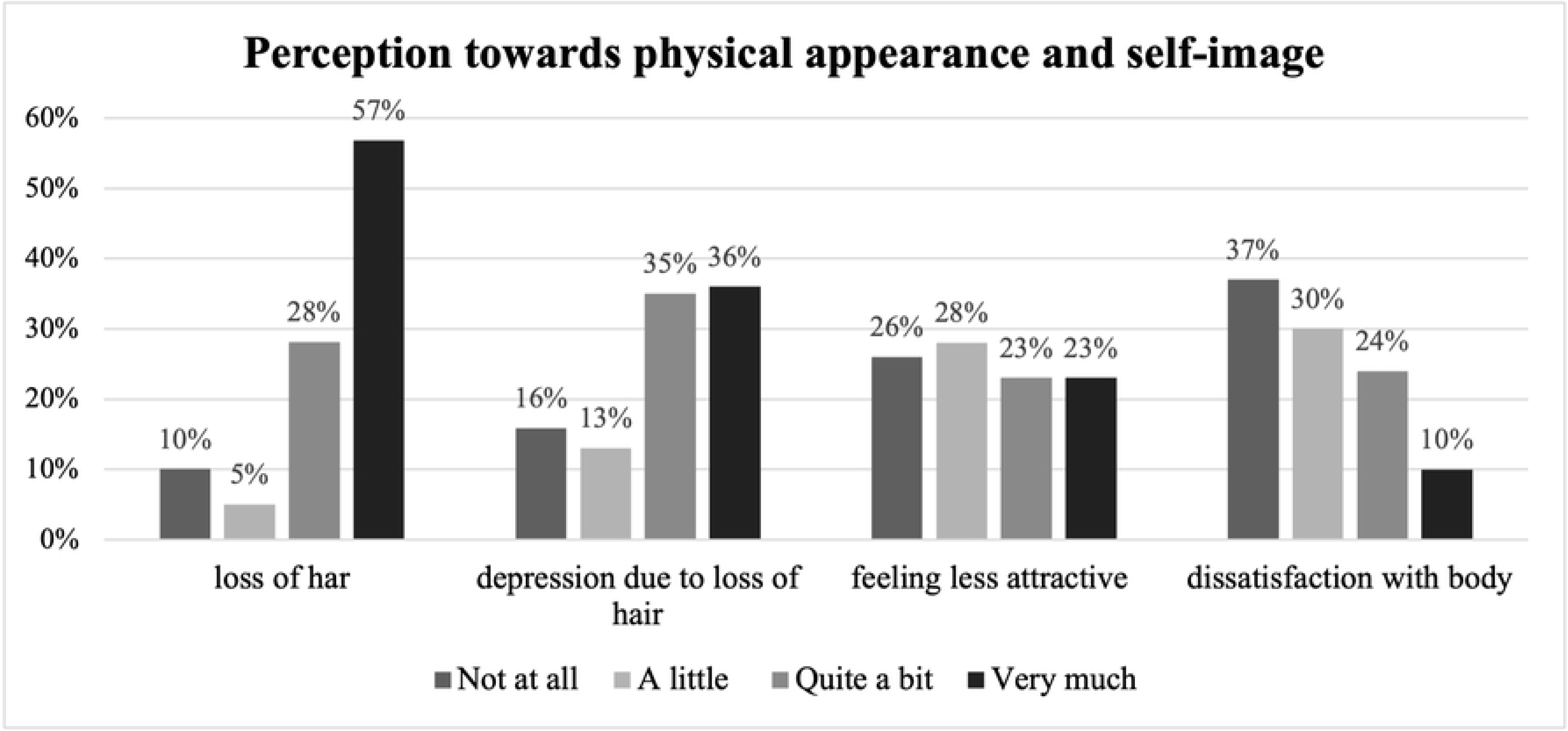
Perception towards physical appearance and self-image of breast cancer patients.

70% of our respondents claimed that they were “not at all” dissatisfied with their sexual activities with their partners despite feeling less attractive or unsatisfied with their self-image. **Fig 7** shows the reaction.

**Fig 7.**
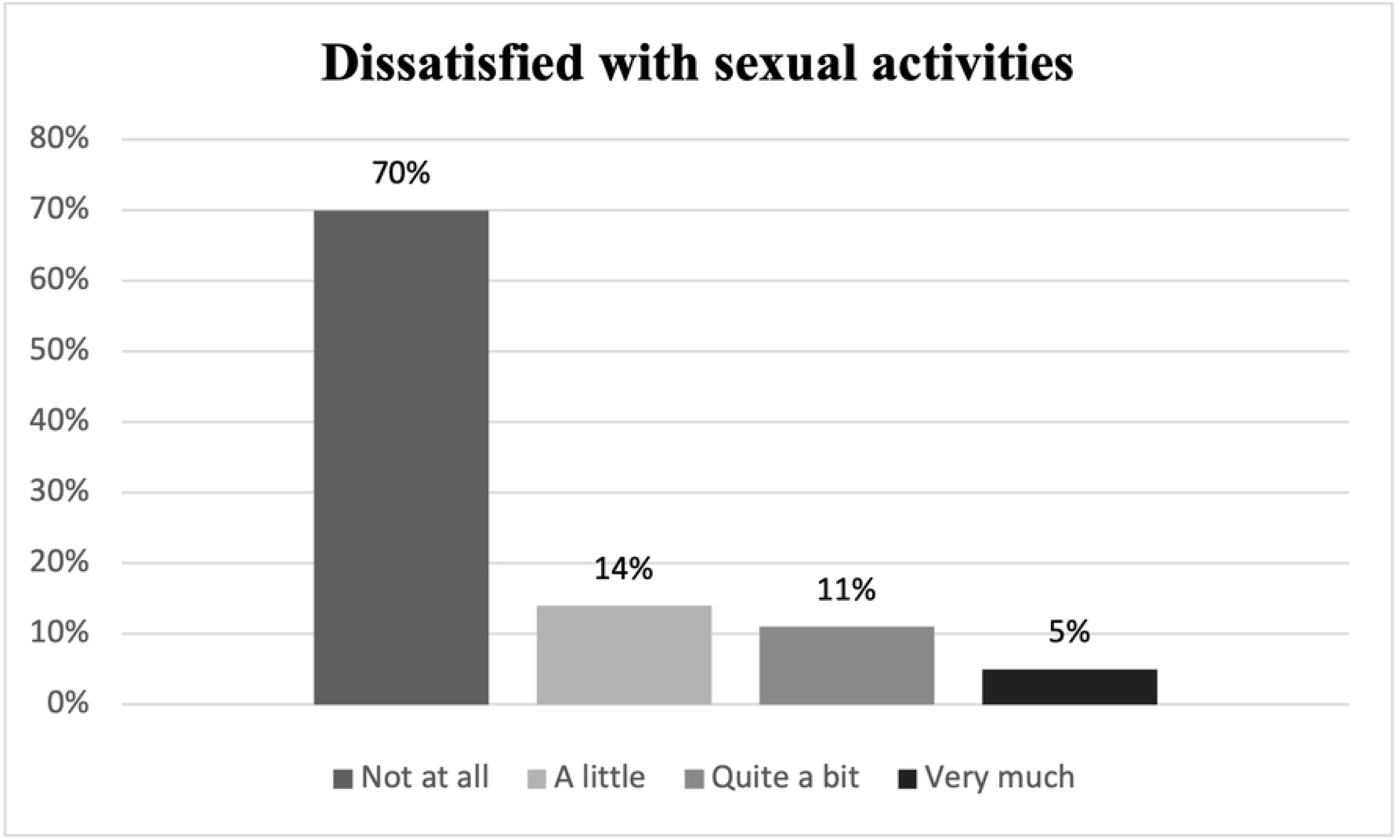
Dissatisfied with sexual activities.

### Quality of life in breast cancer survivors

Data from quality of life assessment by using EORTC QLQ-C30 questionnaires are mentioned in **Table 3**. Overall for EORTC-C30, high scores in global health status and function and function scales indicate good quality of life. On the contrary, high scores in symptoms scale, represent the poor quality of life. Among the 359 respondents, global health status was poor in 28.14 % as they scored <33.3. For functional scale, cognitive and social functioning were high as 42.90% and 38.44% of the respondent scored > 66.7. While for symptom scale the score was moderate-to-low for most of the items. Financial difficulties (28.97%) and fatigue (17.82%) were the most disturbing factors reported by them followed by loss of appetite as 14.76% and 13.65%. Diarrhoea was the least disturbing symptom, as only 2.51% scored >66.7.

**Table 3.** Quality of life in breast cancer survivors by using EORTC QLQ-C30.

| Scales | N | No. of items | Mean± SD | 95% CI | scoring< 33.3<br>N(%)* | scoring> 66.7<br>N(%)* |
| --- | --- | --- | --- | --- | --- | --- |
| **QLQ-C30 questionnaire** |  |  |  |  |  |  |
| Global health status/ QoL | 359 | 2 | 42.85±23.96 | [40.36-45.33] | 101(28.14) | 32(8.91) |
| **Functional scales\*\*** |  |  |  |  |  |  |
| Physical functioning | 359 | 5 | 57.64±24.26 | [55.12-60.16] | 52(14.49) | 107(29.81) |
| Role functioning | 359 | 2 | 54.78±27.85 | [51.89-57.67] | 55(15.32) | 69(19.22) |
| Emotional Functioning | 359 | 4 | 51.88±25.33 | [49.25-54.50] | 75(20.90) | 103(28.69) |
| Cognitive functioning | 359 | 2 | 65.32±27.58 | [62.46-68.18] | 31(8.64) | 154(42.90) |
| Social functioning | 359 | 2 | 66.57±28.97 | [63.57-69.58] | 35(9.75) | 138(38.44) |
| **Symptom scales\*\*\*** |  |  |  |  |  |  |
| Fatigue | 359 | 3 | 51.25±23.13 | [48.85-53.65] | 39(10.86) | 64(17.82) |
| Nausea and vomiting | 359 | 2 | 30.97±24.75 | [28.40-33.53] | 161(44.85) | 17(4.46) |
| Pain | 359 | 2 | 44.29±25.36 | [41.66-46.92] | 79(22.01) | 43(11.98) |
| Dyspnoea | 359 | 1 | 41.23±31.66 | [37.94-44.51] | 89(24.79) | 40(11.14) |
| Insomnia | 359 | 1 | 45.96±32.98 | [42.54-49.38] | 84(23.40) | 49(13.65) |
| Appetite | 359 | 1 | 49.66±32.28 | [46.32-53.03] | 70(19.50) | 53(14.76) |
| Constipation | 359 | 1 | 31.94±34.99 | [28.31-35.57] | 170(47.35) | 35(9.75) |
| Diarrhoea | 359 | 1 | 12.53±25.54 | [9.88-15.19] | 278(77.44) | 9(2.51) |
| Financial difficulties | 359 | 1 | 70.00±25.43 | [67.37-72.65] | 20(5.57) | 104(28.97) |

[\* For functional scales, subjects scoring < 33.3% have a poor quality of life; those scoring 66.7% have a good quality of life
\* For symptom scales/symptoms, subjects scoring < 33.3% have a good quality of life; those scoring = 66.7% have a poor quality of life
\*\* For functional scales, higher scores indicate better functioning.
\*\*\* For symptom scales, higher scores indicate worse functioning]

### Association between dependent and independent variables

Univariate and multivariable logistic regression was conducted to see the association of socio-demographic characteristics (age, religion, region, education level, occupation, marital status, family structure, years diagnosed with cancer & menstrual status) with the outcome variable. We calculated both unadjusted and adjusted odds ratios with 95% CI.

### Global Health Status

**Table 4** presents logistic regression analyses which determined whether the independent variables affected global health status. The model predicts that holding the effects of all other variables constant, it is expected that female patients with breast cancer from the Muslim religion have lower odds of having a good quality of life, in comparison to those who were from other religions (AOR 0.22, p=0.012). However, religion showed no statistically significant relationship with poor quality of life as the p-value is more than 0.05.

**Table 4.** Multiple logistic regression table for association of Global health status with socio-demographic characteristics.

| Global health scale/ QOL | scoring < 33.3<br>N (%) |  |  |  |  | scoring > 66.7<br>N (%) |  |  |  |  |
| --- | --- | --- | --- | --- | --- | --- | --- | --- | --- | --- |
| Characteristics | N (%) | UOR<br>[95% CI] | P<br>value | AOR<br>[95% CI] | P<br>value | N (%) | UOR<br>[95% CI] | P<br>value | AOR<br>[95% CI] | P<br>value |
| **Age (years)** |  |  |  |  |  |  |  |  |  |  |
| Less than 40 years | 48<br>(47.52) | Ref |  | Ref |  | 15<br>(46.88) | Ref |  | Ref |  |
| 40-49 years | 27<br>(26.73) | 0.99 [0.56-1.76] | 0.972 | 0.94 [0.50-1.76] | 0.847 | 7<br>(21.88) | 0.82 [0.32-2.14] | 0.686 | 1.04 [0.36-2.96] | 0.944 |
| More than 50 years | 26<br>(25.74) | 0.67 [0.38-1.17] | 0.161 | 0.59 [0.30-1.14] | 0.116 | 10<br>(31.25) | 0.82 [0.35-1.93] | 0.654 | 0.78 [0.27-2.24] | 0.644 |
| **Religion** |  |  |  |  |  |  |  |  |  |  |
| Muslim | 95<br>(94.06) | 0.97 [0.36-2.62] | 0.946 | 0.95 [0.34-2.64] | 0.928 | 27<br>(84.38) | 0.33 [0.11-1.00] | 0.049 | 0.22 [0.07-0.72] | 0.012 |
| Others | 6<br>(5.94) | Ref |  | Ref |  | 5<br>(15.63) | Ref |  | Ref |  |
| **Region** |  |  |  |  |  |  |  |  |  |  |
| Dhaka | 53<br>(52.48) | 1.05 [0.65-1.67] | 0.848 | 1.06 [0.66-1.73] | 0.801 | 12<br>(37.5) | 0.57 [0.27-1.22] | 0.147 | 0.50 [0.22-1.12] | 0.092 |
| Outside Dhaka | 48<br>(47.52) | Ref |  | Ref |  | 20<br>(62.5) | Ref |  | Ref |  |
| **Education level** |  |  |  |  |  |  |  |  |  |  |
| No education | 41<br>(40.59) | **Ref** |  | **Ref** |  | 10<br>(31.25) | **Ref** |  | **Ref** |  |
| Primary education | 28<br>(27.72) | 0.88 [0.50-1.55] | 0.65<br>2 | 0.85 [0.47-1.54] | 0.591 | 11<br>(34.38) | 1.41 [0.57-3.50] | 0.454 | 1.54 [0.58-4.11] | 0.388 |
| Secondary | 32<br>(31.68) | 1.55 [0.87-2.74] | 0.13<br>7 | 1.54 [0.83-2.85] | 0.172 | 11<br>(34.38) | 2.18 [0.87-5.47] | 0.098 | 2.06 [0.75-5.64] | 0.158 |
| **Employment status** |  |  |  |  |  |  |  |  |  |  |
| Housewife | 87<br>(86.14) | 0.84 [0.42-1.69] | 0.62<br>9 | 0.94 [0.45-1.97] | 0.872 | 24<br>(75) | 0.41 [0.17-1.00] | 0.049 | 0.55 [0.20-1.52] | 0.249 |
| Others | 14<br>(13.86) | **Ref** |  | **Ref** |  | 8<br>(25) | **Ref** |  | **Ref** |  |
| **Marital status** |  |  |  |  |  |  |  |  |  |  |
| Married | 85<br>(84.16) | 1.07 [0.57-2.03] | 0.82<br>7 | 0.93 [0.45-1.89] | 0.831 | 21<br>(65.63) | 0.39 [0.17-0.87] | 0.021 | 0.32 [0.12-0.85] | 0.023 |
| Others | 16<br>(15.84) | **Ref** |  | **Ref** |  | 11<br>(34.38) | **Ref** |  | **Ref** |  |
| **Family structure** |  |  |  |  |  |  |  |  |  |  |
| Single-family | 81<br>(80.2) | 1.33 [0.75-2.37] | 0.32<br>6 | 1.30 [0.72-2.34] | 0.384 | 22<br>(68.75) | 0.72 [0.32-1.62] | 0.434 | 0.75 [0.32-1.76] | 0.506 |
| Joint family | 20<br>(19.8) | **Ref** |  | **Ref** |  | 10<br>(31.25) | **Ref** |  | **Ref** |  |
| **Years first diagnosed with cancer** |  |  |  |  |  |  |  |  |  |  |
| Less than 1 years | 67<br>(66.34) | **Ref** |  | **Ref** |  | 20<br>(62.5) | **Ref** |  | **Ref** |  |
| 1-2 years | 19<br>(18.81) | 1.39 [0.74-2.63] | 0.30<br>9 | 1.59 [0.81-3.10] | 0.178 | 4<br>(12.5) | 0.98 [0.31-3.06] | 0.974 | 1.14 [0.33-3.92] | 0.833 |
| More than 2 years | 15<br>(14.85) | 0.95 [0.49-1.85] | 0.88<br>0 | 0.97 [0.49-1.92] | 0.934 | 8<br>(25) | 1.70 [0.69-4.15] | 0.246 | 1.89 [0.71-5.02] | 0.204 |
| **Menstruation status** |  |  |  |  |  |  |  |  |  |  |
| Menstruating | 25<br>(24.75) | 0.91 [0.53-1.56] | 0.73<br>2 | 0.74 [0.40-1.36] | 0.333 | 11<br>(34.38) | 1.45 [0.66-3.18] | 0.355 | 1.18 [0.47-2.97] | 0.731 |
| Amenorrhea /<br>menopause | 76<br>(75.25) | **Ref** |  | **Ref** |  | 21<br>(65.63) | **Ref** |  | **Ref** |  |
|  |  |  | **Cons** | 0.47 [0.10-2.18] | 0.333 |  |  | **Cons** | 2.20 [0.31-15.68] | 0.432 |

The model also shows that, when the effects of all other factors remain constant, it is expected that, married women have lower odds of having a good quality of life compared to those women who were currently not married (AOR 0.32, P=0.023). However, marital status has no significant association with having poor quality of life. Additionally, employment status as a housewife was significant in unadjusted analyses but was not found significant after adjusting for other factors.

### Physical functioning scales

The logistic model presented in the table showed in **Table 5** predicts that holding the effects of all other factors constant, it is expected that odds of having poor physical functioning quality of life is higher among female patients with breast cancer from an age group more than 50 years than those who were from age group less than 40 years (AOR 3.59, p=0.005). However, age showed no statistically significant relationship with good physically functioning quality of life.

**Table 5.**
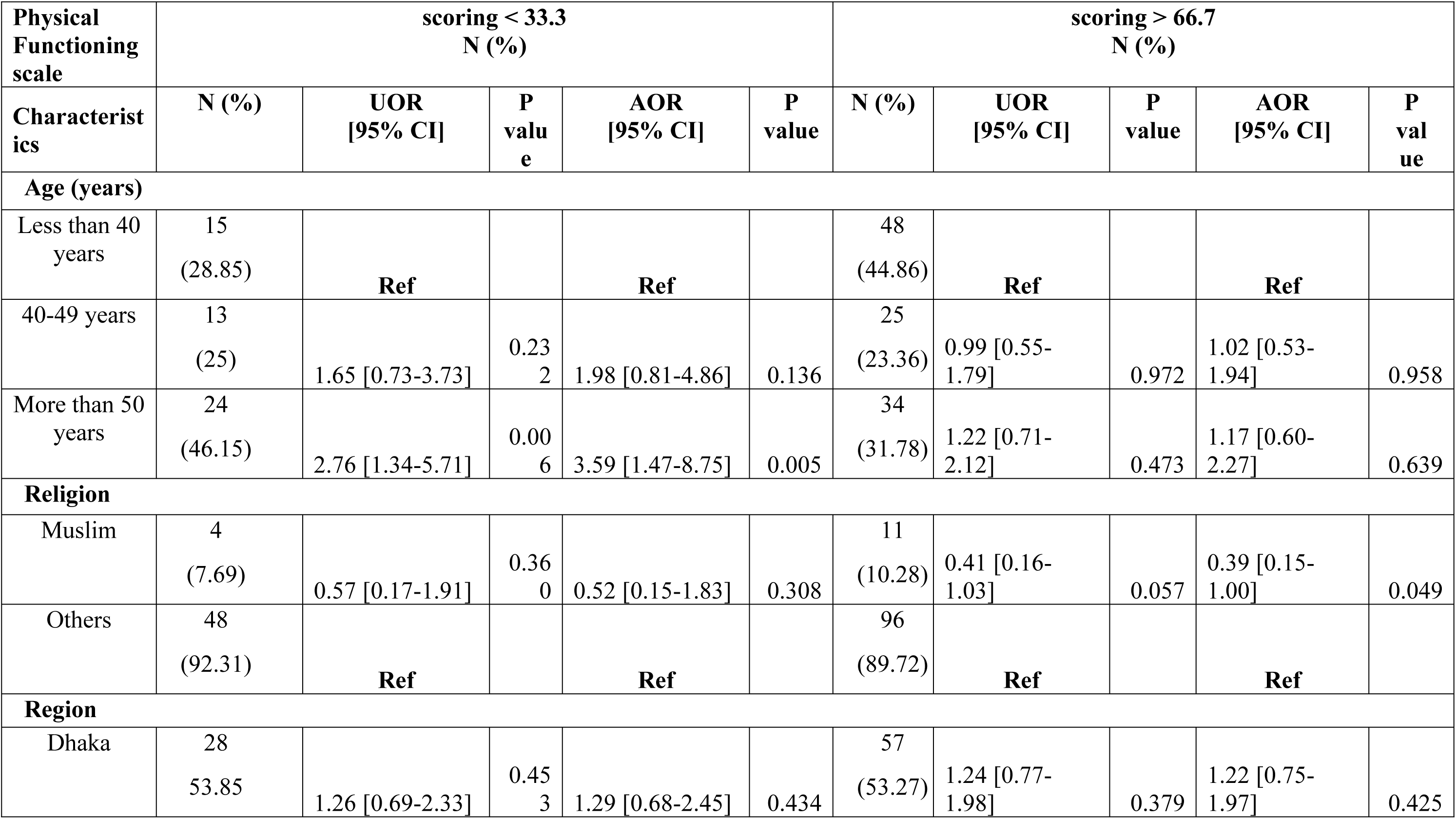
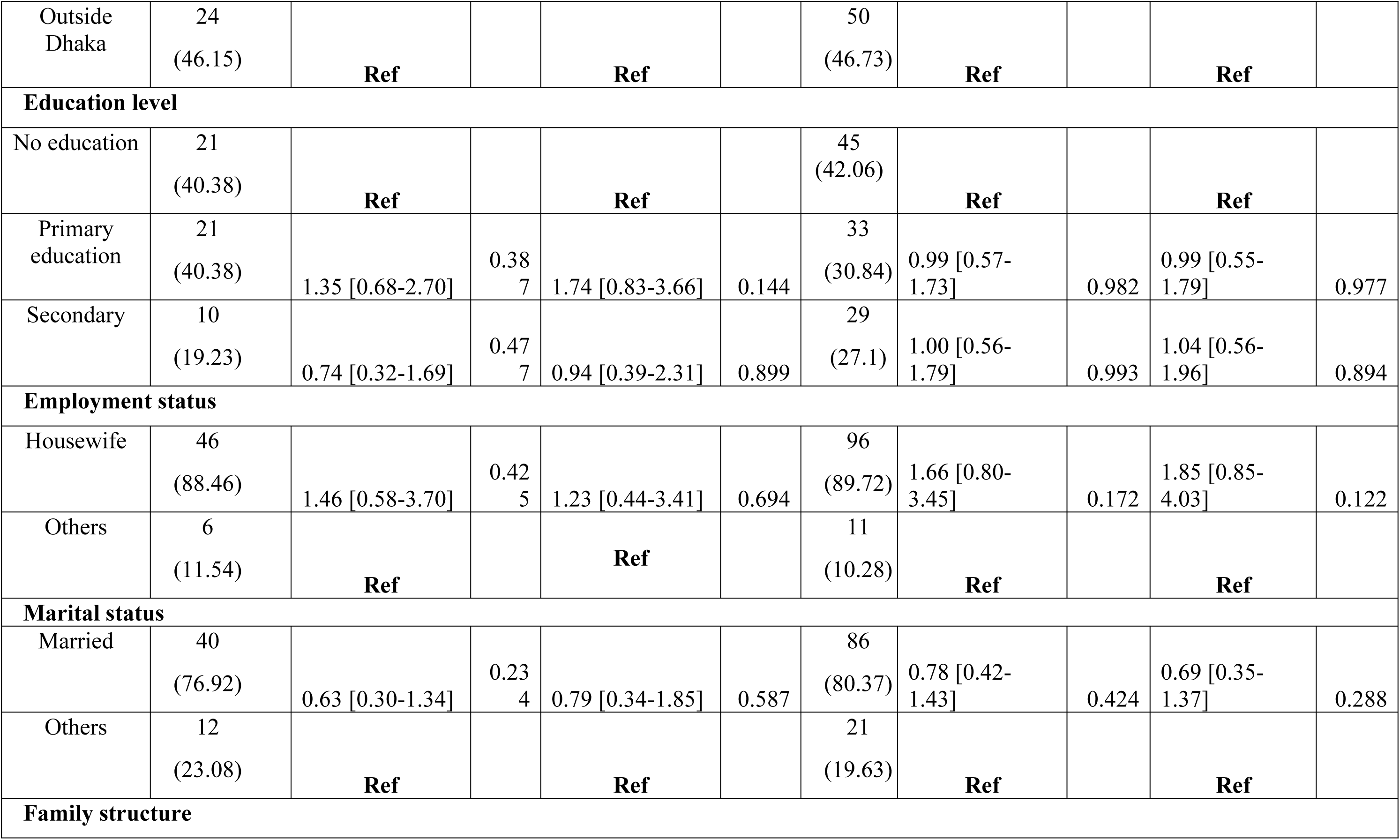
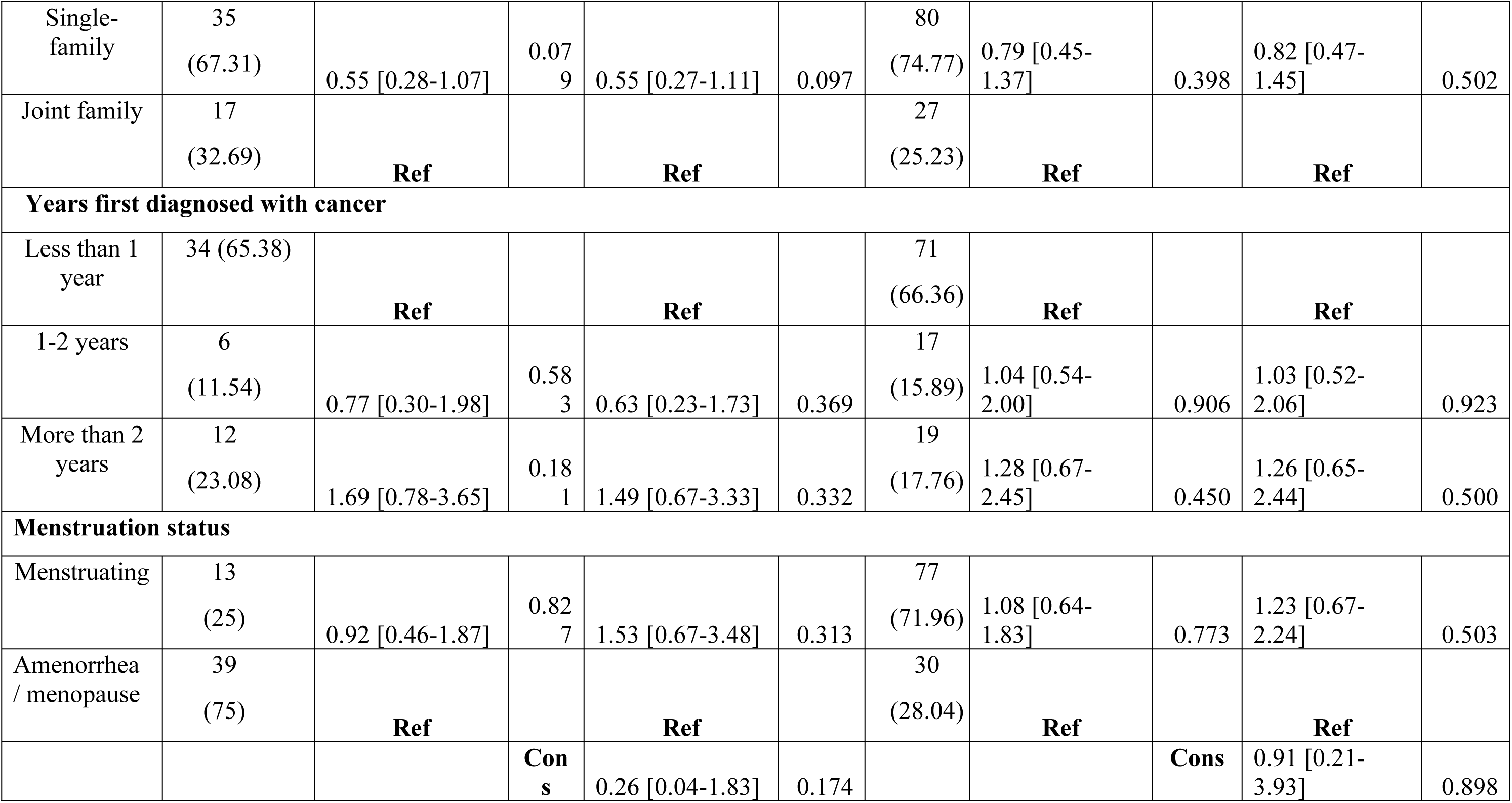
Multiple logistic regression model: for association of Physical functioning scale with socio-demographic characteristics.

Another prediction from the model is that, when the effects of all other factors remain constant, it is expected that odds of experiencing good physically functioning life is lower among those patients who follow the Muslim religion than who were from other religion (AOR 0.39, p=0.049).

### Emotional functioning scale

Details of the model are shown in the table of **Table 6** The model predicts that the odds of having poor emotionally functioning quality of life was higher in female patients from age group 40-49 years compared to the patients from age group less than 40 years (AOR=3.17, p=0.003), holding the effects of other factors constant and this result is statistically significant. This model also predicts that if other factors remain constant, women from an age group more than 50 years had higher odds of having a poor emotionally functioning quality of life than women between 40-49 years (AOR 2.87, p=0.0

**Table 6.** Multiple logistic regression model: for association of Emotional functioning scale with socio-demographic characteristics.

| Emotional<br>Functioning scale | scoring < 33.3<br>N (%) |  |  |  |  | scoring > 66.7<br>N (%) |  |  |  |  |
| --- | --- | --- | --- | --- | --- | --- | --- | --- | --- | --- |
| Characteristics | N (%) | UOR<br>[95% CI] | P<br>valu<br>e | AOR<br>[95% CI] | P<br>valu<br>e | N (%) | UOR<br>[95% CI] | P<br>value | AOR<br>[95% CI] | P<br>val<br>ue |
| **Age (years)** |  |  |  |  |  |  |  |  |  |  |
| Less than 40 years | 20<br>(26.67<br>) | **Ref** |  | **Ref** |  | 53<br>(51.46) | **Ref** |  | **Ref** |  |
| 40-49 years | 27<br>(36) | 2.87 [1.44-5.72] | 0.00<br>3 | 3.17 [1.49-6.78] | 0.00<br>3 | 21<br>(20.39) | 0.84 [0.45-1.58] | 0.592 | 0.94 [0.48-1.87] | 0.866 |
| More than 50 years | 28<br>(37.33) | 2.13 [1.09-4.13] | 0.02<br>6 | 2.87 [1.30-6.35] | 0.00<br>9 | 29<br>(28.16) | 0.83 [0.47-1.46] | 0.519 | 1.23 [0.62-2.42] | 0.555 |
| **Religion** |  |  |  |  |  |  |  |  |  |  |
| Muslim | 69<br>(92) | 0.74 [0.26-2.09] | 0.57<br>5 | 0.75 [0.25-2.23] | 0.60<br>1 | 96<br>(93.2) | 0.89 [0.33-2.36] | 0.81 | 1.02 [0.37-2.78] | 0.977 |
| Others | 6<br>(8) | **Ref** |  | **Ref** |  | 7<br>(6.8) | **Ref** |  | **Ref** |  |
| **Region** |  |  |  |  |  |  |  |  |  |  |
| Dhaka | 40<br>(53.33) | 1.26 [0.74-2.16] | 0.39<br>7 | 1.21 [0.68-2.13] | 0.51<br>4 | 55<br>(53.4) | 1.27 [0.78-2.06] | 0.341 | 1.26 [0.76-2.07] | 0.371 |
| Outside Dhaka | 35<br>(46.67) | **Ref** |  | **Ref** |  | 48<br>(46.6) | **Ref** |  | **Ref** |  |
| **Education level** |  |  |  |  |  |  |  |  |  |  |
| No education | 32<br>(42.67<br>) | **Ref** |  | **Ref** |  | 43<br>(41.75) | **Ref** |  | **Ref** |  |
| Primary education | 28 | 1.26 [0.68-2.34] | 0.46 | 1.64 [0.84-3.21] | 0.14 | 36 | 1.21 [0.69-2.13] | 0.514 | 1.09 [0.59- | 0.790 |
|  | (37.33) |  | 1 |  | 6 | (34.95) |  |  | 1.99] |  |
| Secondary | 15<br>(20) | 0.65 [0.32-1.32] | 0.23<br>3 | 0.73 [0.34-1.57] | 0.42<br>3 | 24<br>(23.3) | 0.78 [0.42-1.43] | 0.413 | 0.60 [0.31-1.16] | 0.130 |
| **Employment status** |  |  |  |  |  |  |  |  |  |  |
| Housewife | 65<br>(86.67) | 0.85 [0.38-1.91] | 0.69<br>9 | 0.48 [0.19-1.18] | 0.10<br>8 | 85<br>(82.52) | 0.62 [0.31-1.23] | 0.169 | 0.46 [0.22-0.97] | 0.041 |
| Others | 10<br>(13.33) | **Ref** |  | **Ref** |  | 18<br>(17.48) | **Ref** |  | **Ref** |  |
| **Marital status** |  |  |  |  |  |  |  |  |  |  |
| Married | 62<br>(82.67) | 1.31 [0.65-2.62] | 0.44<br>7 | 1.89 [0.85-4.20] | 0.12<br>0 | 90<br>(87.38) | 1.90 [0.96-3.76] | 0.064 | 2.53 [1.18-5.44] | 0.018 |
| Others | 13<br>(17.33) | **Ref** |  | **Ref** |  | 13<br>(12.62) | **Ref** |  | **Ref** |  |
| **Family structure** |  |  |  |  |  |  |  |  |  |  |
| Single-family | 53<br>(70.67) | 0.66 [0.36-1.22] | 0.18<br>5 | 0.59 [0.31-1.13] | 0.10<br>9 | 78<br>(75.73) | 0.86 [0.48-1.52] | 0.597 | 0.78 [0.43-1.41] | 0.408 |
| Joint family | 22<br>(29.33) | **Ref** |  | **Ref** |  | 25<br>(24.27) | **Ref** |  | **Ref** |  |
| **Years first diagnosed with cancer** |  |  |  |  |  |  |  |  |  |  |
| Less than 1 year | 49<br>(65.33) | **Ref** |  | **Ref** |  | 74<br>(71.84) | **Ref** |  | **Ref** |  |
| 1-2 years | 8<br>(10.67) | 0.58 [0.25-1.34] | 0.20<br>4 | 0.55 [0.22-1.33] | 0.18<br>2 | 13<br>(12.62) | 0.63 [0.31-1.26] | 0.189 | 0.68 [0.32-1.43] | 0.309 |
| More than 2 years | 18<br>(24) | 1.71 [0.86-3.40] | 0.12<br>6 | 1.64 [0.80-3.37] | 0.18<br>1 | 16<br>(15.53) | 1.01 [0.51-2.00] | 0.986 | 1.14 [0.56-2.34] | 0.710 |
| **Menstruation status** |  |  |  |  |  |  |  |  |  |  |
| Menstruating | 15 | 0.71 [0.37-1.37] | 0.31 | 0.99 [0.47-2.11] | 0.98 | 34 | 1.40 [0.83-2.38] | 0.207 | 1.39 [0.76- | 0.287 |
|  | (20) |  | 2 |  | 7 | (33.01) |  |  | 2.55] |  |
| Amenorrhea/<br>menopause | 60<br>(80) | **Ref** |  | **Ref** |  | 69<br>(66.69) | **Ref** |  | **Ref** |  |
|  |  |  | **Cons** | 0.38 [0.07-2.12] | 0.26<br>9 |  |  | **Cons** | 0.55 [0.12-2.52] | 0.441 |

This model also predicts that, when the effect of other factors held constant, odds of having a good emotionally functioning quality of life is lower (AOR 0.46, p=0.041) in women who were housewife compared to those from other occupational status. Another prediction from the model is that when the effects of other factors are constant odds of having good emotionally functioning quality of life is higher in women who are married at present than those who are not (AOR 2.53, p=0.018).

### Social functioning scale

**Table 7** shows that the model predicts odds of having poor social functioning and quality of life is 5.23 times higher in women from age group more than 50 years than those who are from an age group less than 40 years when effects of other factors remain constant and the relationship is statistically significant (p=0.003).

**Table 7.** Multiple logistic regression model: for association of Social functioning scale with socio-demographic characteristic.

| **Social Functioning scale** | **scoring < 33.3**<br>**N (%)** |  |  |  |  | **scoring > 66.7**<br>**N (%)** |  |  |  |  |
| --- | --- | --- | --- | --- | --- | --- | --- | --- | --- | --- |
| **Characteristics** | **N (%)** | **UOR**<br>**[95% CI]** | **P**<br>**valu**<br>**e** | **AOR**<br>**[95% CI]** | **P**<br>**valu**<br>**e** | **N (%)** | **UOR**<br>**[95% CI]** | **P**<br>**value** | **AOR**<br>**[95% CI]** | **P**<br>**val**<br>**ue** |
| **Age (years)** |  |  |  |  |  |  |  |  |  |  |
| Less than 40 years | 8<br>(22.86) | **Ref** |  | **Ref** |  | 61<br>(44.2) | **Ref** |  | **Ref** |  |
| 40-49 years | 9<br>(25.71) | 2.09 [0.76-5.76] | 0.156 | 2.27 [0.77-6.74] | 0.139 | 31<br>(22.46) | 0.94 [0.54-1.64] | 0.834 | 1.08 [0.59-1.99] | 0.803 |
| More than 50 years | 18<br>(51.43) | 4.09 [1.66-10.08] | 0.002 | 5.23 [1.78-15.34] | 0.003 | 46<br>(33.33) | 1.37 [0.82-2.30] | 0.234 | 2.01 [1.07-3.77] | 0.030 |
| **Religion** |  |  |  |  |  |  |  |  |  |  |
| Muslim | 33<br>(94.29) | 1.34 [0.29-6.19] | 0.705 | 1.44 [0.30-7.01] | 0.653 | 130<br>(94.2) | 1.32 [0.54-3.25] | 0.542 | 1.42 [0.56-3.58] | 0.458 |
| Others | 2 (5.71) | **Ref** |  | **Ref** |  | 8 (5.8) | **Ref** |  | **Ref** |  |
| **Region** |  |  |  |  |  |  |  |  |  |  |
| Dhaka | 18<br>(51.43) | 1.04 [0.50-2.13] | 0.923 | 1.08 [0.51-2.28] | 0.849 | 69<br>(50) | 0.98 [0.63-1.52] | 0.924 | 1.00 [0.63-1.59] | 0.988 |
| Outside Dhaka | 17<br>(48.57) | **Ref** |  | **Ref** |  | 69<br>(50) | **Ref** |  | **Ref** |  |
| **Education level** |  |  |  |  |  |  |  |  |  |  |
| No education | 16<br>(45.71) | **Ref** |  | **Ref** |  | 52<br>(37.68) | **Ref** |  | **Ref** |  |
| Primary education | 10<br>(28.57) | 0.81 [0.35-1.91] | 0.636 | 1.12 [0.45-2.77] | 0.801 | 43<br>(31.16) | 1.08 [0.64-1.81] | 0.782 | 1.15 [0.66-2.01] | 0.617 |
| Secondary | 9<br>(25.71) | 1.13 [0.46-2.76] | 0.79<br>7 | 1.73 [0.65-4.63] | 0.27<br>1 | 43<br>(31.16) | 1.65 [0.95-2.87] | 0.074 | 1.70 [0.93-3.11] | 0.083 |
| **Employment status** |  |  |  |  |  |  |  |  |  |  |
| Housewife | 30<br>(85.71) | 0.85 [0.30-2.40] | 0.75<br>4 | 0.78 [0.25-2.47] | 0.67<br>6 | 117<br>(84.78) | 0.79 [0.42-1.49] | 0.459 | 0.69 [0.34-1.38] | 0.293 |
| Others | 5<br>(14.29) | **Ref** |  | **Ref** |  | 21<br>(15.22) | **Ref** |  | **Ref** |  |
| **Marital status** |  |  |  |  |  |  |  |  |  |  |
| Married | 26<br>(74.29) | 0.65 [0.28-1.50] | 0.31<br>1 | 0.98 [0.37-2.54] | 0.96<br>0 | 116<br>(84.06) | 1.18 [0.65-2.12] | 0.582 | 1.61 [0.83-3.12] | 0.161 |
| Others | 9<br>(25.71) | **Ref** |  | **Ref** |  | 22<br>(15.94) | **Ref** |  | **Ref** |  |
| **Family structure** |  |  |  |  |  |  |  |  |  |  |
| Single-family | 27<br>(77.14) | 0.98 [0.42-2.33] | 0.97<br>1 | 1.01 [0.42-2.47] | 0.97<br>5 | 102<br>(73.91) | 0.83 [0.50-1.38] | 0.466 | 0.85 [0.50-1.44] | 0.545 |
| Joint family | 8<br>(22.86) | **Ref** |  | **Ref** |  | 36<br>(26.09) | **Ref** |  | **Ref** |  |
| **Years first diagnosed with cancer** |  |  |  |  |  |  |  |  |  |  |
| Less than 1 year | 26<br>(74.29) | **Ref** |  | **Ref** |  | 88<br>(63.77) | **Ref** |  | **Ref** |  |
| 1-2 years | 5<br>(14.29) | 0.78 [0.28-2.19] | 0.63<br>9 | 0.55 [0.19-1.64] | 0.28<br>6 | 18<br>(13.04) | 0.83 [0.44-1.57] | 0.569 | 0.80 [0.41-1.57] | 0.522 |
| More than 2 years | 4<br>(11.43) | 0.83 [0.27-2.60] | 0.75<br>4 | 0.68 [0.21-2.19] | 0.51<br>7 | 32<br>(23.19) | 1.97 [1.09-3.57] | 0.025 | 2.04 [1.10-3.78] | 0.024 |
| **Menstruation status** |  |  |  |  |  |  |  |  |  |  |
| Menstruating | 6<br>(17.14) | 0.59 [0.23-1.52] | 0.27<br>8 | 0.97 [0.33-2.81] | 0.95<br>3 | 42<br>(30.43) | 1.26 [0.77-2.05] | 0.358 | 1.37 [0.78-2.41] | 0.269 |
| Amenorrhea /<br>menopause | 29<br>(82.86) | **Ref** |  | **Ref** |  | 96<br>(69.57) | **Ref** |  | **Ref** |  |
|  |  |  | **Cons** | 0.07 [0.01-0.77] | 0.03<br>0 |  |  | **Cons** | 0.31 [0.08-1.26] | 0.102 |

The model predicted that when all other factors are constant the odds of having good social functioning and quality of life are 2.04 times higher in women who were diagnosed with breast cancer more than 2 years compared to women who have been diagnosed for less than 1 year and it statistically significant (p=0.024)

### Role functioning scale

**Table 8** shows that the model predicts the odds of having poor role functioning quality of life among women from the age group 40-49 years was higher than those who were from the age group less than 40 years (AOR 3.89, p=0.002) when effects of other factors remain constant. The model also predicts that the odds of having poor role functioning quality of life is higher in women fan from an age group more than 50 years than those who are from an age group less than 40 years when effects of other factors remain constant (AOR 3.14, p=0.013).

**Table 8.**
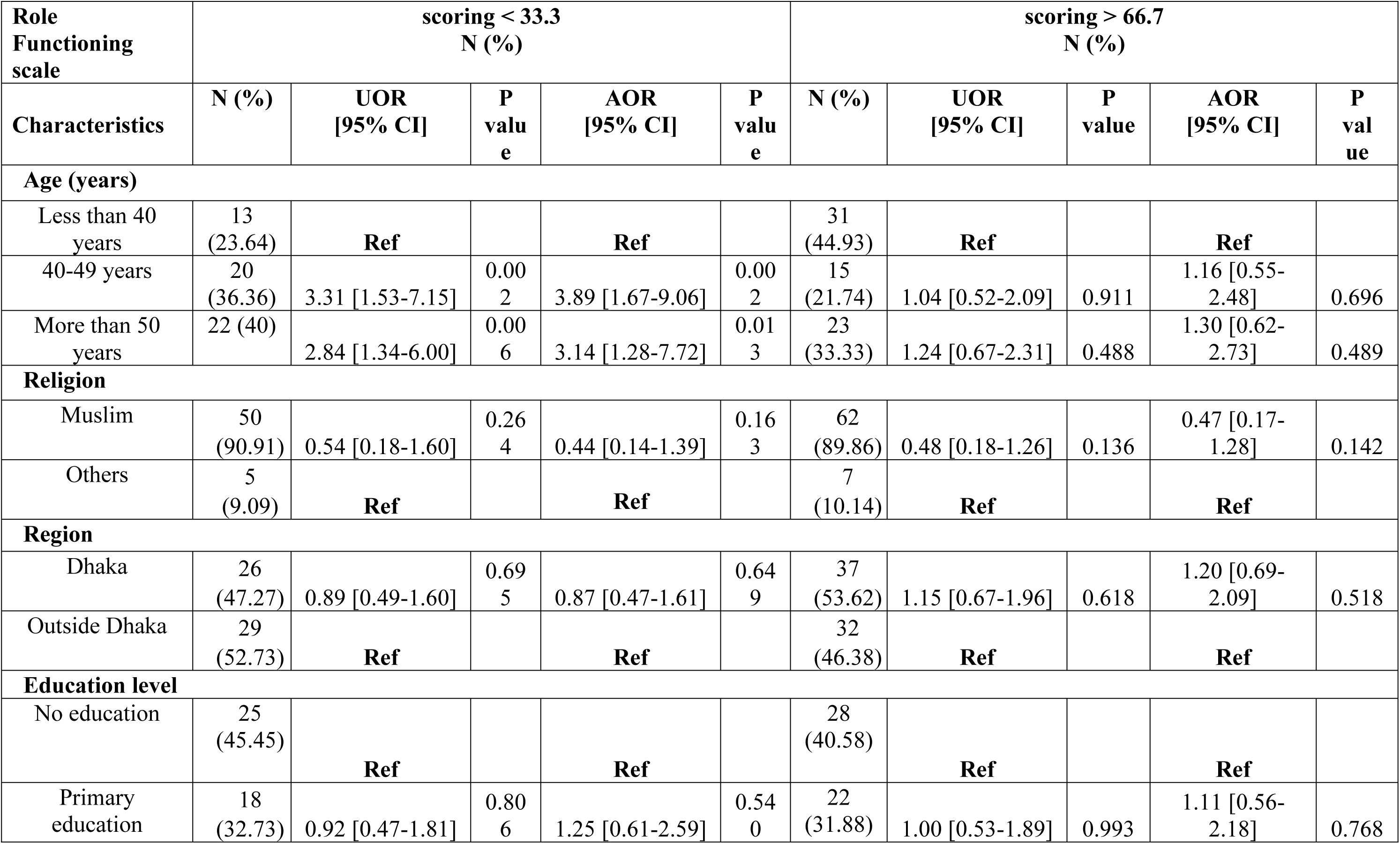
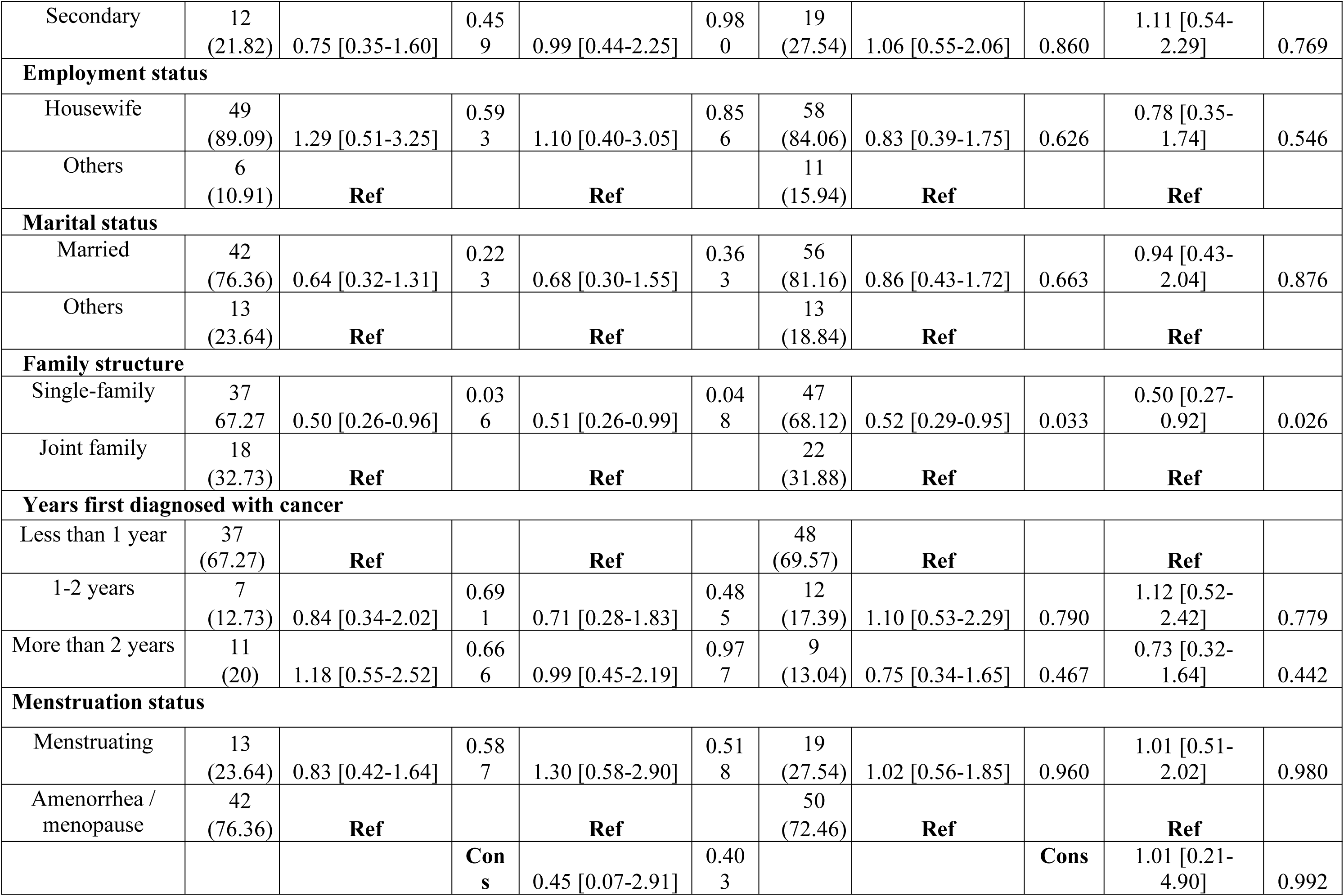
Multiple logistic regression model: for association of Role functioning scale with socio-demographic characteristics.

| **Role**<br>**Functioning**<br>**scale** | **scoring < 33.3**<br>**N (%)** |  |  |  |  | **scoring > 66.7**<br>**N (%)** |  |  |  |  |
| --- | --- | --- | --- | --- | --- | --- | --- | --- | --- | --- |
| **Characteristics** | **N (%)** | **UOR**<br>**[95% CI]** | **P**<br>**valu**<br>**e** | **AOR**<br>**[95% CI]** | **P**<br>**valu**<br>**e** | **N (%)** | **UOR**<br>**[95% CI]** | **P**<br>**value** | **AOR**<br>**[95% CI]** | **P**<br>**val**<br>**ue** |
| **Age (years)** |  |  |  |  |  |  |  |  |  |  |
| Less than 40 years | 13<br>(23.64) | **Ref** |  | **Ref** |  | 31<br>(44.93) | **Ref** |  | **Ref** |  |
| 40-49 years | 20<br>(36.36) | 3.31 [1.53-7.15] | 0.00<br>2 | 3.89 [1.67-9.06] | 0.00<br>2 | 15<br>(21.74) | 1.04 [0.52-2.09] | 0.911 | 1.16 [0.55-2.48] | 0.696 |
| More than 50 years | 22 (40) | 2.84 [1.34-6.00] | 0.00<br>6 | 3.14 [1.28-7.72] | 0.01<br>3 | 23<br>(33.33) | 1.24 [0.67-2.31] | 0.488 | 1.30 [0.62-2.73] | 0.489 |
| **Religion** |  |  |  |  |  |  |  |  |  |  |
| Muslim | 50<br>(90.91) | 0.54 [0.18-1.60] | 0.26<br>4 | 0.44 [0.14-1.39] | 0.16<br>3 | 62<br>(89.86) | 0.48 [0.18-1.26] | 0.136 | 0.47 [0.17-1.28] | 0.142 |
| Others | 5<br>(9.09) | **Ref** |  | **Ref** |  | 7<br>(10.14) | **Ref** |  | **Ref** |  |
| **Region** |  |  |  |  |  |  |  |  |  |  |
| Dhaka | 26<br>(47.27) | 0.89 [0.49-1.60] | 0.69<br>5 | 0.87 [0.47-1.61] | 0.64<br>9 | 37<br>(53.62) | 1.15 [0.67-1.96] | 0.618 | 1.20 [0.69-2.09] | 0.518 |
| Outside Dhaka | 29<br>(52.73) | **Ref** |  | **Ref** |  | 32<br>(46.38) | **Ref** |  | **Ref** |  |
| **Education level** |  |  |  |  |  |  |  |  |  |  |
| No education | 25<br>(45.45) | **Ref** |  | **Ref** |  | 28<br>(40.58) | **Ref** |  | **Ref** |  |
| Primary education | 18<br>(32.73) | 0.92 [0.47-1.81] | 0.80<br>6 | 1.25 [0.61-2.59] | 0.54<br>0 | 22<br>(31.88) | 1.00 [0.53-1.89] | 0.993 | 1.11 [0.56-2.18] | 0.768 |
| Secondary | 12<br>(21.82) | 0.75 [0.35-1.60] | 0.45<br>9 | 0.99 [0.44-2.25] | 0.98<br>0 | 19<br>(27.54) | 1.06 [0.55-2.06] | 0.860 | 1.11 [0.54-2.29] | 0.769 |
| **Employment status** |  |  |  |  |  |  |  |  |  |  |
| Housewife | 49<br>(89.09) | 1.29 [0.51-3.25] | 0.59<br>3 | 1.10 [0.40-3.05] | 0.85<br>6 | 58<br>(84.06) | 0.83 [0.39-1.75] | 0.626 | 0.78 [0.35-1.74] | 0.546 |
| Others | 6<br>(10.91) | **Ref** |  | **Ref** |  | 11<br>(15.94) | **Ref** |  | **Ref** |  |
| **Marital status** |  |  |  |  |  |  |  |  |  |  |
| Married | 42<br>(76.36) | 0.64 [0.32-1.31] | 0.22<br>3 | 0.68 [0.30-1.55] | 0.36<br>3 | 56<br>(81.16) | 0.86 [0.43-1.72] | 0.663 | 0.94 [0.43-2.04] | 0.876 |
| Others | 13<br>(23.64) | **Ref** |  | **Ref** |  | 13<br>(18.84) | **Ref** |  | **Ref** |  |
| **Family structure** |  |  |  |  |  |  |  |  |  |  |
| Single-family | 37<br>67.27 | 0.50 [0.26-0.96] | 0.03<br>6 | 0.51 [0.26-0.99] | 0.04<br>8 | 47<br>(68.12) | 0.52 [0.29-0.95] | 0.033 | 0.50 [0.27-0.92] | 0.026 |
| Joint family | 18<br>(32.73) | **Ref** |  | **Ref** |  | 22<br>(31.88) | **Ref** |  | **Ref** |  |
| **Years first diagnosed with cancer** |  |  |  |  |  |  |  |  |  |  |
| Less than 1 year | 37<br>(67.27) | **Ref** |  | **Ref** |  | 48<br>(69.57) | **Ref** |  | **Ref** |  |
| 1-2 years | 7<br>(12.73) | 0.84 [0.34-2.02] | 0.69<br>1 | 0.71 [0.28-1.83] | 0.48<br>5 | 12<br>(17.39) | 1.10 [0.53-2.29] | 0.790 | 1.12 [0.52-2.42] | 0.779 |
| More than 2 years | 11<br>(20) | 1.18 [0.55-2.52] | 0.66<br>6 | 0.99 [0.45-2.19] | 0.97<br>7 | 9<br>(13.04) | 0.75 [0.34-1.65] | 0.467 | 0.73 [0.32-1.64] | 0.442 |
| **Menstruation status** |  |  |  |  |  |  |  |  |  |  |
| Menstruating | 13<br>(23.64) | 0.83 [0.42-1.64] | 0.58<br>7 | 1.30 [0.58-2.90] | 0.51<br>8 | 19<br>(27.54) | 1.02 [0.56-1.85] | 0.960 | 1.01 [0.51-2.02] | 0.980 |
| Amenorrhea /<br>menopause | 42<br>(76.36) | **Ref** |  | **Ref** |  | 50<br>(72.46) | **Ref** |  | **Ref** |  |
|  |  |  | **Cons** | 0.45 [0.07-2.91] | 0.40<br>3 |  |  | **Cons** | 1.01 [0.21-4.90] | 0.992 |

The model also predicts lower odds of having poor role functioning quality of life in women who live in single-family than those who were from a joint family when the effects of other factors remain constant (AOR 0.51, p=0.048). The model also predicts the odds of having a good role functioning quality of life is lower (AOR 0.50, p=0.026) in women who live in single-family than those who were from a joint family when effects of other factors remain constant and the relationship is statistically significant

### Cognitive functioning scale

Details of the model are given in **Table 9**. The model predicts the odds of having a poor cognitive functioning quality of life is lower in women who came from Dhaka compared to those women who came from outside Dhaka division when effects of other factors remain constant (AOR 0.48, p=0.007).

**Table 9.** Multiple logistic regression model: for association of Cognitive functioning scale with socio-demographic characteristics.

| **Cognitive Functioning scale** | **scoring < 33.3**<br>**N (%)** |  |  |  |  | **scoring > 66.7**<br>**N (%)** |  |  |  |  |
| --- | --- | --- | --- | --- | --- | --- | --- | --- | --- | --- |
| **Characteristics** | **N (%)** | **UOR**<br>**[95% CI]** | **P**<br>**valu**<br>**e** | **AOR**<br>**[95% CI]** | **P**<br>**valu**<br>**e** | **N (%)** | **UOR**<br>**[95% CI]** | **P**<br>**value** | **AOR**<br>**[95% CI]** | **P**<br>**val**<br>**ue** |
| **Age (years)** |  |  |  |  |  |  |  |  |  |  |
| Less than 40 years | 14<br>(45.16) | **Ref** |  | **Ref** |  | 70<br>(45.45) | **Ref** |  | **Ref** |  |
| 40-49 years | 8<br>(25.81) | 0.96 [0.37-2.47] | 0.93<br>4 | 1.08 [0.39-3.05] | 0.88<br>0 | 36<br>(23.38) | 0.86 [0.50-1.50] | 0.604 | 0.99 [0.55-1.79] | 0.973 |
| More than 50 years | 9<br>(29.03) | 0.85 [0.34-2.10] | 0.72<br>4 | 1.30 [0.44-3.83] | 0.63<br>3 | 48<br>(31.17) | 0.91 [0.55-1.50] | 0.702 | 1.28 [0.70-2.35] | 0.417 |
| **Religion** |  |  |  |  |  |  |  |  |  |  |
| Muslim | 29<br>(93.55) | 0.98 [0.21-4.65] | 0.97<br>8 | 1.02 [0.21-5.04] | 0.98<br>2 | 143<br>(92.86) | 0.88 [0.37-2.08] | 0.767 | 0.99 [0.41-2.41] | 0.986 |
| Others | 2<br>(6.45) | **Ref** |  | **Ref** |  | 11<br>(7.14) | **Ref** |  | **Ref** |  |
| **Region** |  |  |  |  |  |  |  |  |  |  |
| Dhaka | 12<br>(38.71) | 0.53 [0.24-1.15] | 0.10<br>6 | 0.48 [0.21-1.09] | 0.07<br>8 | 74<br>(48.05) | 0.77 [0.50-1.19] | 0.237 | 0.78 [0.50-1.22] | 0.274 |
| Outside Dhaka | 19<br>(61.29) | **Ref** |  | **Ref** |  | 80<br>(51.95) | **Ref** |  | **Ref** |  |
| **Education level** |  |  |  |  |  |  |  |  |  |  |
| No education | 11<br>(38.48) | **Ref** |  | **Ref** |  | 60<br>(38.96) | **Ref** |  | **Ref** |  |
| Primary education | 13<br>(41.94) | 1.87 [0.78-4.49] | 0.16<br>3 | 1.72 [0.68-4.34] | 0.25<br>4 | 53<br>(34.42) | 1.40 [0.84-2.33] | 0.202 | 1.33 [0.77-2.29] | 0.300 |
| Secondary education | 7<br>(22.58) | 1.12 [0.40-3.09] | 0.83<br>1 | 0.88 [0.30-2.60] | 0.81<br>5 | 41<br>(26.62) | 1.20 [0.70-2.06] | 0.509 | 1.09 [0.61-1.95] | 0.767 |
| **Employment status** |  |  |  |  |  |  |  |  |  |  |
| Housewife | 28<br>(90.32) | 1.64 [0.46-5.79] | 0.44<br>2 | 1.18 [0.30-4.64] | 0.81<br>2 | 134<br>(87.01) | 1.18 [0.63-2.21] | 0.611 | 1.03 [0.52-2.03] | 0.929 |
| Others | 3<br>(9.68) | **Ref** |  | **Ref** |  | 20<br>(12.99) | **Ref** |  | **Ref** |  |
| **Marital status** |  |  |  |  |  |  |  |  |  |  |
| Married | 27<br>(87.1) | 2.01 [0.67-6.10] | 0.21<br>5 | 2.03 [0.59-6.99] | 0.26<br>1 | 133<br>(86.36) | 1.89 [1.06-3.38] | 0.031 | 1.89 [1.00-3.60] | 0.051 |
| Others | 4<br>(12.9) | **Ref** |  | **Ref** |  | 21<br>13.64 | **Ref** |  | **Ref** |  |
| **Family structure** |  |  |  |  |  |  |  |  |  |  |
| Single-family | 21<br>(67.74) | 0.67 [0.29-1.53] | 0.34<br>1 | 0.70 [0.30-1.65] | 0.41<br>2 | 120<br>(77.92) | 1.12 [0.68-1.88] | 0.659 | 1.12 [0.66-1.90] | 0.682 |
| Joint family | 10<br>(32.26) | **Ref** |  | **Ref** |  | 34<br>22.08 | **Ref** |  | **Ref** |  |
| **Years first diagnosed with cancer** |  |  |  |  |  |  |  |  |  |  |
| Less than 1 year | 22<br>(70.97) | **Ref** |  | **Ref** |  | 108<br>(70.13) | **Ref** |  | **Ref** |  |
| 1-2 years | 1<br>(3.23) | 0.17 [0.02-1.33] | 0.09<br>2 | 0.18 [0.02-1.44] | 0.10<br>6 | 24<br>(15.58) | 0.84 [0.46-1.54] | 0.579 | 0.89 [0.47-1.66] | 0.706 |
| More than 2 years | 8<br>(25.81) | 1.38 [0.56-3.41] | 0.48<br>3 | 1.63 [0.63-4.24] | 0.31<br>4 | 22<br>(14.29) | 0.77 [0.42-1.42] | 0.410 | 0.86 [0.46-1.60] | 0.628 |
| **Menstruation status** |  |  |  |  |  |  |  |  |  |  |
| Menstruating | 10<br>(32.26) | 1.54 [0.67-3.54] | 0.30<br>5 | 1.48 [0.57-3.82] | 0.41<br>8 | 45<br>(29.22) | 1.34 [0.82-2.19] | 0.246 | 1.32 [0.76-2.31] | 0.330 |
| Amenorrhea / menopause | 21<br>(67.74) | **Ref** |  | **Ref** |  | 109<br>(70.78) | **Ref** |  | **Ref** |  |
|  |  |  | **Cons** | 0.11 [0.01-1.31] | 0.08<br>1 |  |  | **Cons** | 0.43 [0.11-1.69] | 0.228 |

The model also predicts the odds of having a good cognitive functioning quality of life are higher in women who were married compared to those women who were currently not when effects of other factors remain constant (AOR 1.89, P=0.051).

## 5. Discussion

In our study, we categorized the HRQOL score and compared results by using mean scores and the proportion of patients who met the cut-off level of ≥66% for good functioning and ≤33% for poor functioning taking the reference from earlier studies conducted by (8). By using this cut-off score, we found that only 8.91% of the respondents perceived good quality of health. These scores indicated the poor quality of life in our patients. However, these findings contradict other studies conducted in the Saudi Arab (39), Nepal (17), and Bahrain (40). The study conducted in Saudi Arabia with a good sample size (n=284) showed a higher proportion of global health status, with 46% of respondents scoring >66.7. The study from Nepal also showed a higher global health status. A possible reason could be; unlike the mentioned countries, the overall health system of Bangladesh is not well structured and the government of Bangladesh is not prioritizing breast cancer patients enough. In our study, married women from the Muslim religion showed lower odds of experiencing (AOR 0.22, p=0.012) good quality of health in global health status but we could not find any study with the best of our knowledge to compare our result with.

In the cognitive functioning and social functioning scale, 42.90% and 38.44% of our respondents showed higher scores, respectively. This indicated a good quality of functioning life. Among the five domains of functioning scales, only a few patients scored higher on the role functioning scale (around 19.22%). Our study presented that women who live in a single or nuclear family showed lower odds of poor quality of life in the role functioning domain (AOR 0.51, p=0.048). The predetermined roles of men and women in our society, culture, and traditions may be the reason for this finding. In this society, males are expected to provide for their families and take care of the home. Women are essential to the upkeep of the family, the kids, and the community; thus, when they are diagnosed with breast cancer, it limits their ability to fulfil their tasks due to the uncertainty of survival, disfigurement, and incapacity. Their failure to perform their customary duty and their concern for their family may have contributed to this poor role function. This finding is supported by the study conducted in the Saudi Arab (39) where the patients included in that study scored least on the role functioning scale among five domains as well. Another study from Nepal reflected a similar finding (17). This highlights the significance of interventions that target psychological counselling and physical therapy sessions for breast cancer patients as well as their partners and family members.

The patients included in our study were undergoing treatment for breast cancer like conservative surgery, and chemotherapy and 68% of the respondents had been diagnosed less than a year. However, they demonstrated considerably higher scores in physical and cognitive well-being. This finding was considerably different from a study conducted in India, which found very low scores in general physical well-being (36). Bangladesh, as the neighbouring country to India, was expected to achieve similar results, but unlike us, they have considered only patients from post-operative periods which may be the cause of inconsistent results. The postoperative period is usually associated with pain and limited mobility of patients.

The well-being of the family and conjugal life of a woman is often threatened by the occurrence of breast cancer. Increased provision of social support at that time is the crucial factor (16). One study from Malaysia (41) stated, perceived positive support from society, husbands/ partners, and children of breast cancer patients contributes to enhancing the quality of life outcomes. Our cross-sectional study revealed women suffering from breast cancer showed a higher score in social functioning with a mean score of 66.57 (standard deviation 28.97). While we compared the social functional scales with socio-demographic characteristics, it was found that women who aged more than 50 years showed higher odds of having a good social functioning quality of health (AOR 5.23, p=0.030). It also revealed that women who were diagnosed with breast cancer more than 2 years ago had higher odds of having a better social functioning quality of life (AOR 2.04, P=024). It is in alignment with the results of studies from the Bahrain (40) and the Saudi Arab (39). This result might signify the social support our patients receive from their families and society.

Almost all studies suggested anxiety, depression and low emotional functional ability affect the psychological well-being of breast cancer patients which ultimately results in impaired QOL (42). Diagnosis of cancer, fear, and concern about the recurrence of disease and death; disfigurement, loss of hair, perception of self-image, and changes in femininity, appeal, and sexuality, are factors that can cause unexplained emotional torment even years after detection and treatment. Among the 359 women interviewed, 37% of the respondents stated they felt depressed because of the loss of their hair. However, as per the score of the emotional functioning scale, 29% of the patients scored >66.7. This indicated good emotional functioning. This unique finding was also similar to the finding from the Saudi Arab study (39). The explanation might be debatable as it is subjectively related to the social values of our society that females are not very conscious about their appearance in public. The social support from the family indicated above may be the cause of the patients with breast cancer paying less attention to their physical appearance. According to one study, young breast cancer patients exhibit poor emotional functioning because they are more preoccupied with their physical attractiveness (14). Our research, however, did not find any connection between emotional functioning and age

Breast cancer is considered a disease that is related to women’s identity(16). For a healthy reproductive life, active sexual function is an important issue and for breast cancer patients it is even more important. A systematic review pointed out that, disrupted sexual function due to cancer treatment-chemotherapy, total mastectomy; struggle with partners because of unsatisfactory intercourse is responsible for the poor quality of life (16). However, our study revealed contradictory results where a majority (70%) of the respondents from our study said they were “not at all” dissatisfied with sexual activities with their partners along with the fact that women who were married in our study showed higher odds of emotional functioning quality of health (AOR 2.53, p=0.018). The reason can be, they were not comfortable initiating a discussion about this topic and might prefer to discuss it with their physicians which is also suggested by literature (12). This finding highlights the importance of improving consultation and communication with the patients along with their partners before starting the treatment regimens.

In our study, older patients aged 40 years or above, exhibited higher odds of having poor quality of health in most of the parameters e.g. physical function (AOR 3.59, p=0.005), role function (AOR 3.89, p=0.002) and emotional function (AOR, 2.87, p=0.009) domain. This finding was contradictory to studies conducted in Bahrain (40), and Malaysia (41) where women aged >40 showed the poor quality of health. They mentioned the reason behind this observation was that the younger age-group had other contributing factors for compromised QoL e.g. psychological discomfort, weight gain, lack of physical activity during treatment, and stress about their menopausal-related issues. But findings of our study did not reveal any association between quality of life with menstrual status. This finding is supported by literature revealing that pattern of menopausal symptoms is similar among women with breast cancer and women without it (43). It highlights that there should be proper psychosocial counselling that draws attention to the fact that women with breast cancer should not compromise their quality of life with menstrual issues.

Our study reported high scores in cognitive functioning which also contradicts the general speculation that adults with cancer have a great influence on educational and career decisions and general quality of life due to cognitive deficit (44). The reason behind this contradiction could be that the mentioned study explored patients with 5 years post-diagnosis history who are free from cancer and not undergoing treatment presently. Accordingly, we can conclude that cognitive deficits seem to be seen approximately 2 years after treatment and can persist long after treatment, perhaps indefinitely (45). Our cross-sectional survey showed a significant association between cognitive functioning with marital status (AOR 1.89, p=0.051) but did not reveal any significant association with the duration of their diagnosis of cancer.

In our study women scored low to moderate in symptom scales, with financial difficulties being the highest disturbing factor which is also reaffirmed by the fact that almost 75% of the patients took monetary assistance from relatives, and 37% of the respondents had to sell their assets. This finding is supported by several studies from Nepal (17), and South Korea (11) stating that uncertainty and economic distress because of unemployment, lost wages, assets, financial hardships, inadequate health insurance coverage or inability to pay for healthcare is a great concern for breast cancer patients. When the economic burden rises during regular clinical examination, screening, and oncology follow-up treatment, it also impacts the quality of life in terms of physical and psychological functioning domains (21). It is suggested that support from the community in the form of monetary assistance, financial aid, and advice can alleviate the economic domain of poor quality of life (2, 3, 21, 33). The findings of our study highlight the fact that now it is high time that the government of Bangladesh prioritizes the social protection of breast cancer patients. Given the overall cost of cancer treatment, the commencement of health insurance policy, budgeting for cancer treatment, and strengthening financial allowance to cancer patients should be considered by policymakers.

Our survey findings revealed that in the case of symptoms scales, fatigue, loss of appetite, and insomnia were the distressing symptoms for the respondents right after financial burden. A study conducted in Canada (25)supports this finding saying that almost half of the patients included in that study reported fatigue as a problem. The result is consistent with one study from the Saudi Arab (39). Another study states fatigue is closely attached to depression, physical pain, and sleep disturbance which is also following our findings (18).

Though the findings from the overall quality of life measurement scale used in this study are coherent with other studies, we could not establish any causal association with the type of treatments, age, marital status, depression, and quality of life due to the nature of our study. HRQOL is a dynamic phenomenon. Patients from different age groups and different stages of breast cancer can experience compromised quality of life among different domains. That is why all of the social, economic, physical, mental, and emotional aspects of a breast cancer patient should be considered properly.

## 6. Strengths and Limitations of the study

There have been a few studies that have examined the general quality of life of breast cancer patients, but to our knowledge, no research has been done in Bangladesh that specifically addresses this problem. Additionally, this study included women with various cancer stages from various age groups and geographic locations. This study thoroughly illustrated the total quality of life of a breast cancer patient because HRQOL is not a constant phenomenon; it evolves with age and within several functional domains, including physical-emotional-social. The interpretations of this cross-sectional investigation can be utilized as a preliminary step to further evaluate the QOL with a larger sample size, even though demonstrating any causative direction was beyond the scope of this study. Based on the study’s findings, efficient multi-sectoral approaches can be developed and put into practice right away to reduce the obstacles that stand in the way of a breast cancer patient’s QOL improvement.

This study included several restrictions as well. Due to time and resource limitations, we were unable to reach the desired sample size. The statistical analysis between dependent and independent variables might have been impacted by the short sample size. Due to time constraints, we were only able to perform our study in one public tertiary hospital, therefore the findings may not apply to a larger population. The study had no qualitative component because it was questionnaire-based and it was impossible to have a thorough conversation with patients about specific issues. When asked about their income situation, some individuals remained silent. Furthermore, because the researchers conducted face-to-face interviews to gather the data and because the subject was delicate, this may have affected the participants’ responses. Additionally, as all diagnoses, medical histories, and treatment histories were self-reported, there is a possibility for recollection bias and self-reporting bias. Without using any randomization procedures, the study’s participants were chosen by a convenient sampling method. This can potentially have an impact on the study findings’ internal and external validity and data quality.

## 7. Conclusion and Recommendation

This study was carried out to evaluate the quality of life (QOL) of female breast cancer patients in a tertiary hospital in Bangladesh. Only a small percentage of respondents had high scores on the global health scale, while a larger percentage had high scores on the cognitive and social scales related to the functional areas of QOL scales. Better functioning scores demonstrate people’s optimism and perception of social support during this dreadful condition. Lower scores on the role scales, on the other hand, draw attention to the need for psychological treatment for women as well as their partners, kids, and other family members. The patients identified fatigue and financial issues as the most upsetting factors. The findings of this study emphasize strategies and actions to boost financial support for breast cancer patients as well as to deal with fatigue and pain that should be developed and put into practice.

## Data Availability

N/A

## 8. Acknowledgment

We are grateful to and owe a debt of gratitude to each and every participant for their invaluable time and knowledge, without which the survey would not have been possible. All of the nurses and on-call physicians at the day-care facility of the National Institute of Cancer Research and Hospital are appreciated. I want to express my gratitude to our mentors, Dr. Malay Kanti Mridha and Dr. Ilias Mahmud, for their exceptional supervision and advice throughout the entire study. I also want to thank our research assistants for their assistance with data collection. I’m appreciative to everyone in our group for working well together during the entire study period. In the end. I would especially want to thank BRAC University James P Grant School of Public Health for giving us this opportunity to conduct this study.

## Description of author’s roles

K.F Islam worked in conceptualizing, data collection and data extraction, statistical analysis, and writing original manuscript; A. Awal worked in statistical analysis, preparing tables and figures, and supervising data extraction; F.T. Shaon contributed in data collection and extraction; B. Hossain contributed in data collection and extraction; A. Samson contributed in data collection and extraction; J. S. Momo contributed in data collection and extraction; M. Hasan supported in conceptualizing and designing study method; A. A. M. Hanif supported in conceptualizing and designing study method; I. Mahmud critically supervised study design and conceptualization; M. K. Mridha critically supervised study design and conceptualization, and contributed to manuscript editing, and reviewing.

## List of abbreviations

EORTC: European Organization for Research and Treatment of Cancer
HRQL: Health Related Quality of Life
JPGSPH: James P Grant School of Public Health
LMIC: Lower and middle-income countries
NICRH: National Institute of Cancer Research and Hospital
NCD: Non-communicable disease
OOP: Out of pocket
QOL: Quality of Life Questionnaire Core-30
QOL QC-30: Quality of Life
WHO: World Health Organization

